# Tocilizumab in COVID-19 – A Bayesian reanalysis of RECOVERY

**DOI:** 10.1101/2021.06.15.21258966

**Authors:** Arthur M. Albuquerque, Lucas Tramujas, Lorenzo R. Sewanan, James M. Brophy

## Abstract

**Background:** Randomised Evaluation of COVID-19 Therapy (RECOVERY) demonstrated that tocilizumab reduces mortality in hospitalized COVID-19 patients. However, substantial uncertainty remains whether tocilizumab’s effect is similar across clinically relevant subgroups. Whether this uncertainty can be resolved with Bayesian methods is unknown.

**Design, Setting, Participants, and Interventions:** RECOVERY was a controlled, open-label, platform UK trial that randomized (1:1) 4116 adults with oxygen saturation <92% on room air or receiving oxygen therapy with C-reactive protein ≥75 mg/L to either usual care or tocilizumab plus usual care.

**Main outcome measures:** Mortality and hospital discharge within 28 days.

**Methods:** Using Bayesian methods, we combined RECOVERY with evidence-based priors incorporating previous COVID-19 tocilizumab RCTs. The probability of tocilizumab’s benefit for respiratory support and corticosteroid subgroups and sensitivity analyses were performed with different prior distributions and baseline risks.

**Results:** For all-cause mortality, the posterior probabilities of decreased deaths with tocilizumab were >99% and 19% in patients using and not using corticosteroids, respectively. In patients on simple oxygen only, non-invasive ventilation and invasive mechanical ventilation, the probabilities of decreased mortality were 96%, >99% and 77%, respectively. The probabilities for a clinically significant mortality reduction, as assessed by an absolute risk difference > 3% (number needed to treat ≤ 33), were 77%, 96%, 56%, respectively. Sensitivity analyses highlighted the uncertainty and lack of conclusive evidence for tocilizumab’s effect in patients on invasive mechanical ventilation and those without concurrent corticosteroids. Posterior probabilities of benefit for hospital discharge outcome were high and consistent across most subgroups.

**Conclusions:** In this Bayesian reanalysis, COVID-19 hospitalized patients exposed to corticosteroids or on non-invasive ventilation have a high probability of a clinically meaningful mortality benefit from tocilizumab. Tocilizumab also likely improves discharge from hospital in most subgroups. Future research should further address if patients on invasive mechanical ventilation can also benefit from tocilizumab.

## Introduction

The Randomised Evaluation of COVID-19 Therapy (RECOVERY), a large, open-label, platform, randomized controlled trial (RCT), compared tocilizumab to usual care in patients hospitalized with COVID-19.[1] Results showed a benefit for the primary outcome, mortality reduction at 28 days. Further the authors report that “consistent results were seen in all prespecified subgroups of patients, including those receiving systemic corticosteroids” and “regardless of the amount of respiratory support.” The ability to provide reliable subgroup analyses is of obvious importance in enhancing personalized clinical decision-making.[2]

RECOVERY was designed and analyzed following the standard null hypothesis significance testing (NHST) paradigm. However, the limitations of NHST evaluating RCTs, particularly for subgroup analyses, have been long recognized.[3] Moreover, given that subgroup differences were assessed with low-powered interaction tests, the conclusion of consistent results across all patients may be questioned and could profit from further analyses.[2] Bayesian methods have been proposed as an alternative that may permit additional insights.[4–6] In brief, Bayesian analyses can increase power by incorporating evidence from other RCTs; avoid the nullism, dichotomization, and statistical reification of NHST and directly answer the clinical question of interest: what is the probability of tocilizumab’s effect in each pertinent subgroup?[7, 8] These features may contribute to a more thorough understanding of RCT results, such as those in the RECOVERY trial.

Herein, we re-analyze the tocilizumab data using Bayesian methods with a primary goal of providing an enlightened appreciation of the results and the reliability of specific subgroup conclusions.

## Methods

### RECOVERY

RECOVERY was a multi-center, open-label, platform RCT designed to evaluate different drugs, including tocilizumab, in patients hospitalized with COVID-19. In National Health Service hospitals in the UK, 4,116 adults with oxygen saturation <92% on room air or receiving oxygen therapy and with C-reactive protein ≥ 75 mg/L were randomized to either usual care or tocilizumab plus usual care in a 1:1 ratio. The primary outcome was 28-days all-cause mortality, and the main secondary outcomes were time to discharge from hospital, discharge from hospital within 28 days and, among patients not receiving invasive mechanical ventilation at randomization, a composite outcome of invasive mechanical ventilation or death. RECOVERY was designed, with 90% power and alpha = 0.01, to detect a 5% absolute risk reduction assuming a control mortality risk of at least 25%.

### Main outcomes and subgroups

We analyzed both mortality and discharge from hospital within 28 days (hereafter, mortality and hospital discharge) outcomes, focusing on two clusters of clinically pertinent subgroups. First, we analyzed the effect of tocilizumab as a function of adjunctive therapy, specifically whether corticosteroids were used or not during the follow-up period. Second, we analyzed the effect of tocilizumab as a function of baseline respiratory disease severity as defined by the RECOVERY trialists: simple oxygen only; non-invasive ventilation, including patients on high–flow nasal oxygen, continuous positive airway pressure ventilation or other non–invasive ventilation; and invasive mechanical ventilation at randomization.

### Statistical analyses

Bayesian analysis consists of updating prior beliefs with the current RECOVERY data (likelihood) to form a posterior distribution which allows one to make direct probability statements about the treatment effect by calculating the area under the curve.[8] One can calculate not only the probability of any benefit but also of clinically meaningful benefits. In the Bayesian framework, repeated analyses of the same data are not associated with the well-known multiplicity issues of frequentist analyses.[3] We used medians and 95% highest density intervals, defined by the narrowest interval containing 95% of the probability density function, to describe our posterior distributions.[9]

While the above calculations follow the laws of probability, the most contentious issue of a Bayesian analysis is the choice of priors whose potential subjective nature is often portrayed as its Achilles heel. There are multiple ways of eliciting priors,[10] and to remain objective we used data from other tocilizumab COVID-19 RCTs to create evidence-based priors for each corresponding subgroup. Priors were formed by extracting the number of events and sample sizes from applicable tocilizumab RCTs included in a weekly updated systematic living review.[11] The last update of our analysis was on June 12^*th*^, 2021. Further details about the data extraction process can be found in the Supplementary Material.

Our estimands are expressed as risk differences (RD), chosen for their clinical decision-making utility.[2] We estimated the posterior probabilities of benefit for multiple RD thresholds, including RD >0%, >1%, >2%, and >3%, the later three equivalent to the numbers needed to treat (NNT) of 100, 50, and 33, respectively. We derived RDs from the odds ratio.[12]

Posterior distribution calculations for each subgroup and outcome followed a stepwise approach. First, we pooled data from similar studies with a random-effect meta-analysis using a restricted maximum-likelihood estimator to create evidence-based priors. Next, we estimated the log-odds ratio of the RECOVERY and using conjugate normal analysis combined this with the evidence-based priors to determine the log-odds posterior distributions.[8] Next, we drew and exponentiated 100,000 samples from the posterior distributions to change to an odds ratio scale. Lastly, RDs were derived, where the baseline risk was the RECOVERY control arm for that specific subgroup.[5]

We performed sensitivity analyses to test the robustness of our results. As there is no one “correct” prior, we followed previously published recommendations for prior selections,[8, 13] and tested four different mortality prior distributions: non-informative, skeptical, optimistic, and pessimistic. The high variance for the non-informative prior, despite recognized limitations,[8] allows a posterior distribution dominated by RECOVERY data. The skeptical prior reflects clinical equipoise, promoting shrinkage towards null effect and thereby tempers researcher overenthusiasm. The optimistic prior was chosen to reflect the RECOVERY design projection of a 20% relative risk reduction.[1] Lastly, as proposed by recent recommendations,[13] a pessimistic prior reflecting a belief that tocilizumab could possibly cause harm due to, for example, increased secondary infections, was considered.

Consequently, we chose the following means for our sensitivity priors; i) non-informative and skeptical priors’ mean log-odds = 0 (= log[OR=1]), ii) optimistic prior mean = − 0.223 (= log[OR=0.8]) and iii) pessimistic prior mean = 0.223 (= log[OR=1.25], reciprocal of the optimistic prior). We assumed these log priors to be normally distributed with variance equal to the evidence-based priors’ variance of each respective subgroup. The non-informative prior is an exception, which assigned a variance of 10,000. Regarding the secondary hospital discharge outcome, an odds ratio greater than one means benefit, so while we used the same prior means, their signs were reversed. We used the same evidence-based variances for this secondary outcome.

A final sensitivity analysis involved varying the baseline risk, since a RD limitation is its strong dependence on the baseline risk. As mentioned above, our baseline analysis used the observed RECOVERY control arm risk. To test how our posterior distributions with evidence-based priors vary with different baseline risks, we estimated the risk differences with forty different baseline risks (spanning +-20% change from the original risk) for each subgroup.

We pre-registered our data analysis and extraction protocol before any analyses were performed.[14] We had planned to analyze subgroup outcome data as a function of time from symptom onset but abandoned this due to a lack of objective prior evidence. We added two additional analyses to our original protocol: 1) corticosteroids subgroup analysis,[15] and 2) sensitivity analyses with different baseline risks.[5]

All analyses were conducted in R (R Environment version 4.0.4). The data and full analysis code are available at https://bit.ly/3xnTJSV.

This study was exempt from obtaining formal institutional review board approval and the requirement to obtain informed patient consent because it is secondary research of publicly available data sets.

## Results

### All-cause mortality

There were ten potential RCTs to be incorporated for evidence-based prior distributions,[16–25] but one had high-risk of bias and five failed to report the required subgroup analyses and thus are not included in this analysis.[18, 19, 21–23, 25] RECOVERY mortality results were extracted from the peer reviewed publication,[1] and are presented in Table 1 as a function of baseline disease severity as determined by respiratory status and as a function of adjunctive corticosteroids therapy. Results from our search for prior tocilizumab mortality evidence is also presented in Table 1.[16, 17, 20, 24] The data extraction processes and additional clinical details for each trial are shown in the Supplementary Material.

**Table 1.** Number of events and sample sizes on the mortality outcome. Studies other than RECOVERY represent the data incorporated in the prior distribution for each subgroup.

| Study | Control |  |  | Tocilizumab |  |  |
| --- | --- | --- | --- | --- | --- | --- |
|  | Events | Total | Risk (%) | Events | Total | Risk (%) |
| **Not using corticosteroids** |  |  |  |  |  |  |
| RECOVERY | 127 | 367 | 35 | 139 | 357 | 39 |
| COVACTA | 10 | 65 | 15 | 29 | 188 | 15 |
| REMAP-CAP | 9 | 36 | 25 | 6 | 37 | 16 |
| **Using corticosteroids** |  |  |  |  |  |  |
| RECOVERY | 600 | 1721 | 35 | 482 | 1664 | 29 |
| COVACTA | 18 | 79 | 23 | 29 | 106 | 27 |
| REMAP-CAP | 134 | 361 | 37 | 102 | 358 | 28 |
| **Simple oxygen only** |  |  |  |  |  |  |
| RECOVERY | 214 | 933 | 23 | 180 | 935 | 19 |
| COVACTA | 2 | 44 | 5 | 9 | 78 | 12 |
| CORIMUNO-19 | 8 | 67 | 12 | 7 | 63 | 11 |
| **Non-invasive ventilation** |  |  |  |  |  |  |
| RECOVERY | 366 | 867 | 42 | 310 | 819 | 38 |
| COVACTA | 12 | 39 | 31 | 18 | 94 | 19 |
| REMAP-CAP | 82 | 273 | 30 | 53 | 242 | 22 |
| Salvarani | 1 | 63 | 2 | 2 | 60 | 3 |
| **Invasive mechanical ventilation** |  |  |  |  |  |  |
| RECOVERY | 149 | 294 | 51 | 131 | 268 | 49 |
| COVACTA | 14 | 55 | 25 | 31 | 113 | 27 |
| REMAP-CAP | 60 | 124 | 48 | 45 | 108 | 42 |

Figure 1 and Supplementary Table 1 show the estimated evidence-based prior, RECOVERY data (likelihood), and posterior distributions on tocilizumab’s effect for each sub-group. Although multiple studies are available to compute the prior distribution, RECOVERY provided a notably greater amount of evidence for all subgroups as witnessed by the close approximation of the RECOVERY and the final posterior distributions. As expected, the addition of the RECOVERY data has greatly improved the precision of our prior estimates of tocilizumab’s effect as seen by the narrowing of the posterior distributions. Examining the specific clinical subgroups, the benefit of tocilizumab is especially evident for patients using corticosteroids and those not requiring invasive ventilation.

**Figure 1.**
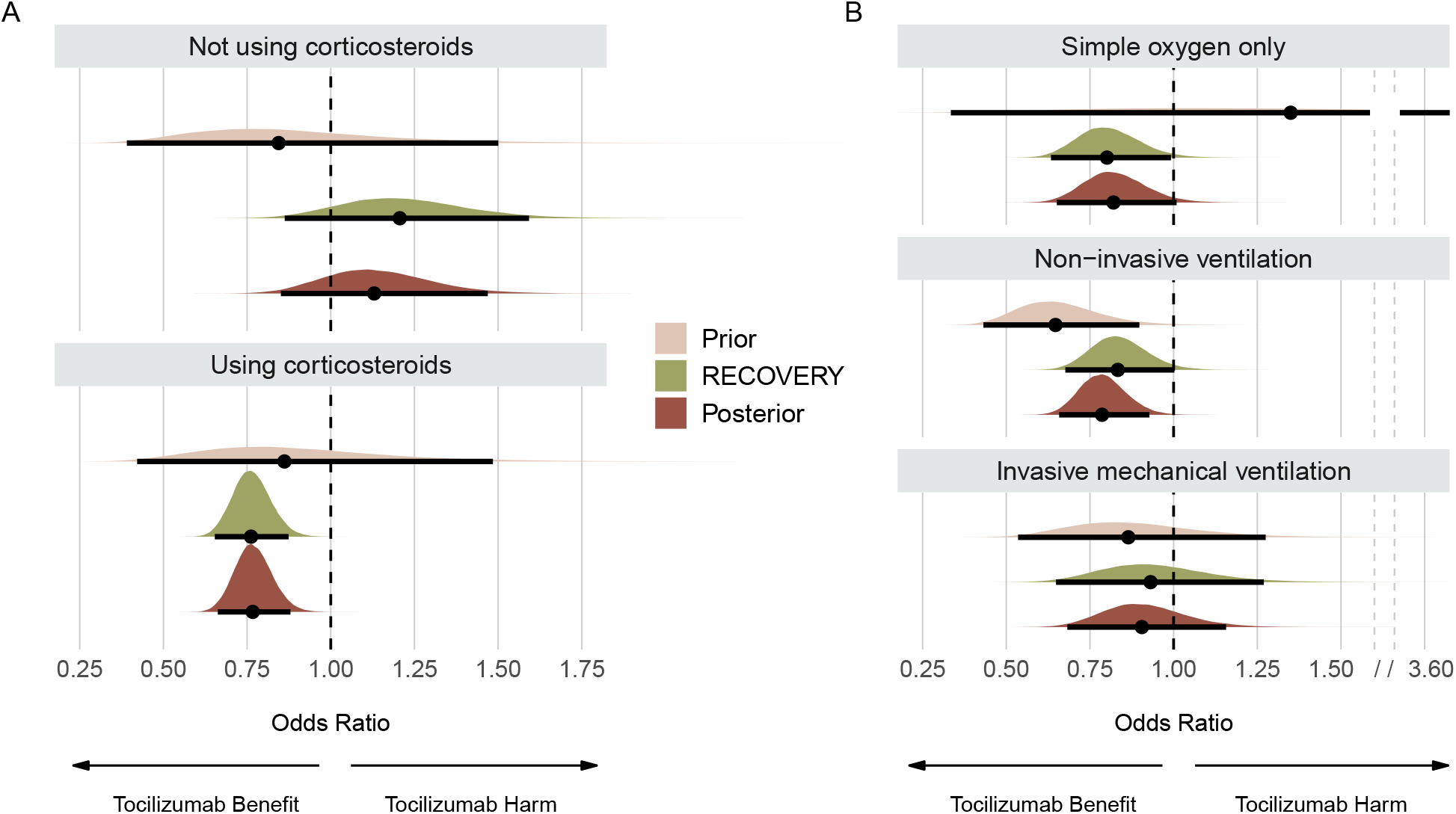
Prior, RECOVERY, and posterior distributions for each subgroup on the mortality outcome. Panel A shows results for subgroups regarding use of corticosteroids. Panel B shows results for subgroups regarding respiratory support. Point estimates depict the median and interval bars depict 95% highest density intervals. Using conjugate normal analyses, these distributions were originally combined in the log-odds ratio scale (Methods section). They were transformed into the odds ratio scale for this figure to aid visual interpretation.

Figure 2 and Supplementary Table 2 show the posterior distributions and probabilities for each subgroup using the evidence-based priors on the risk difference scale. In patients not using corticosteroids, the median risk difference was -2.8 (95% highest density interval [HDI], -9.2 to 3.5) and the posterior probability of a risk difference benefit (≥ 0%) was only 18.9% (Figure 2A shaded dark blue area). In patients using corticosteroids, the median risk difference was 5.8 (95% HDI, 2.8 to 8.6) and the posterior probability of a risk difference benefit was >99% (Figure 2A shaded light blue area). Cumulative posterior probability distributions as a function of varying risk differences are shown in Figure 2B. Importantly, there is an approximately 96% probability that the tocilizumab benefit is at least 3 lives saved (NNT ≤ 33) among patients receiving corticosteroids (Supplementary Table 2).

**Figure 2.**
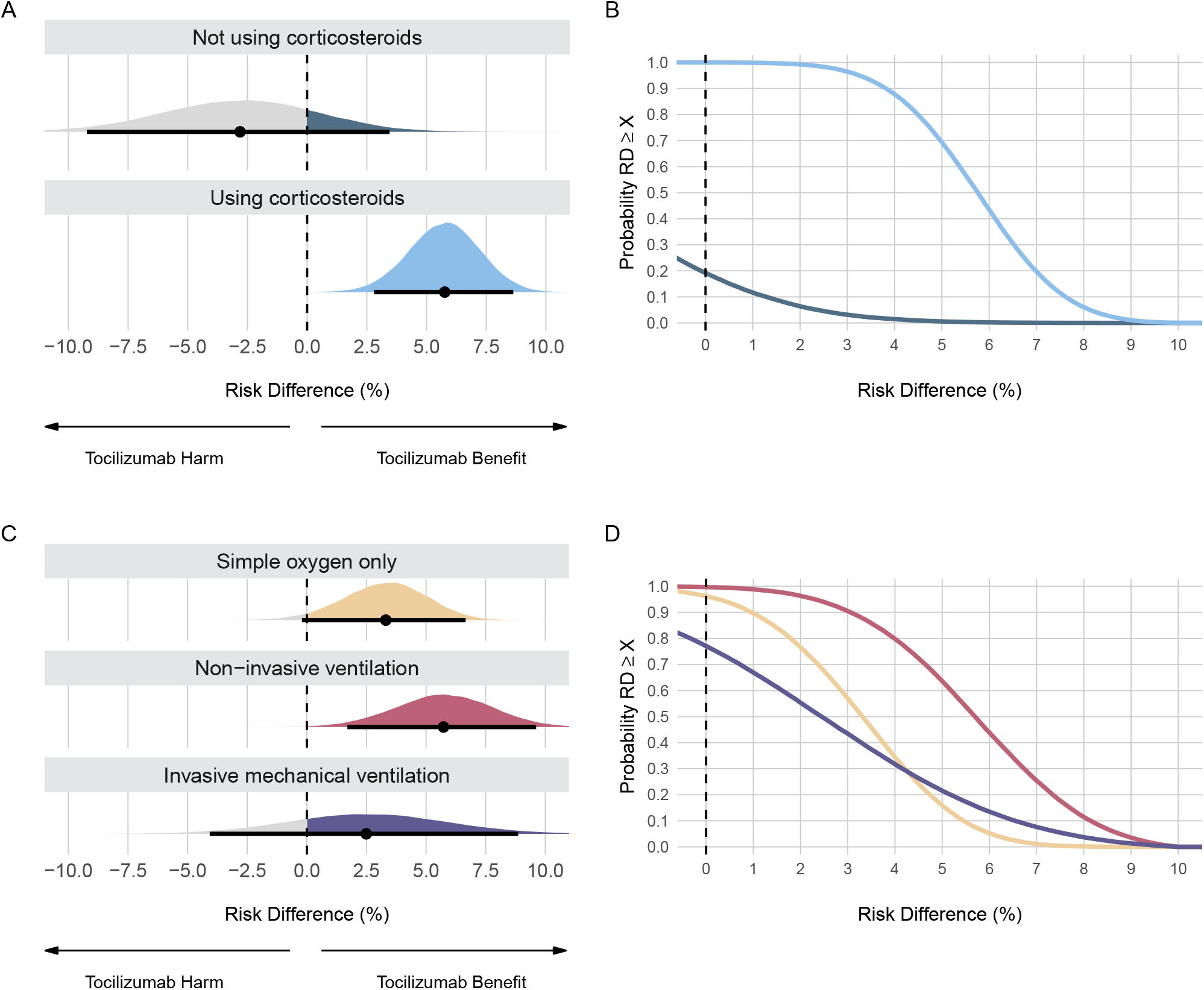
Posterior distributions and probabilities using evidence-based priors on the mortality outcome. Panel A shows the posterior distributions and Panel B shows the cumulative posterior probabilities on subgroups regarding use of corticosteroids. Panel C shows the posterior distributions and Panel D shows the cumulative posterior probabilities on subgroups regarding respiratory support. Panels A and C: Point estimates depict the median and interval bars depict the 95% highest density intervals. Panels B and D: Cumulative posterior distributions correspond to the probabilities that the risk difference (RD) is lower than or equal to the effect size on the X-axis. The colors in Panels B and D match the ones used in Panels A and C.

In patients on simple oxygen only, non-invasive ventilation, and invasive mechanical ventilation, the risk differences were 3.3 (95% HDI, -0.2 to 6.7), 5.7 (95% HDI, 1.7 to 9.6), and 2.5 (95% HDI, -4.1 to 8.9), respectively (Supplementary Table 2). The posterior probabilities of a risk difference benefit were 96.2%, >99%, and 77.3% (Figure 2C). The posterior probabilities for benefits as large as 3% risk difference (NNT ≤ 33) were 57%, 90.6%, and 44% in patients on simple oxygen, non-invasive ventilation, and invasive ventilation, respectively (Figure 2D).

The robustness of Figure 2 results to the choice of priors, graphically displayed in Supplementary Figure 1, is demonstrated graphically in Figure 3 and numerically in Supplementary Table 3. For example, even with a skeptical prior, there was a > 94% probability of tocilizumab benefit for the corticosteroid, simple oxygen, and non-invasive ventilation subgroups. Similarly, the lack of clear tocilizumab benefit for the non-corticosteroid and invasive ventilation subgroups were not substantially modified by the choice of different priors. For example, the posterior probability for any benefit among the invasive mechanical ventilation subgroup ranged from 38% with the pessimistic prior distribution to only 83% with the optimistic prior (Supplementary Table 4). Moreover, the posterior probabilities of a substantial clinical benefit (> 3% risk difference [NNT < 33]) for this subgroup remained modest at 44% for the evidence based and 52.6% for the optimistic prior, respectively.

**Figure 3.**
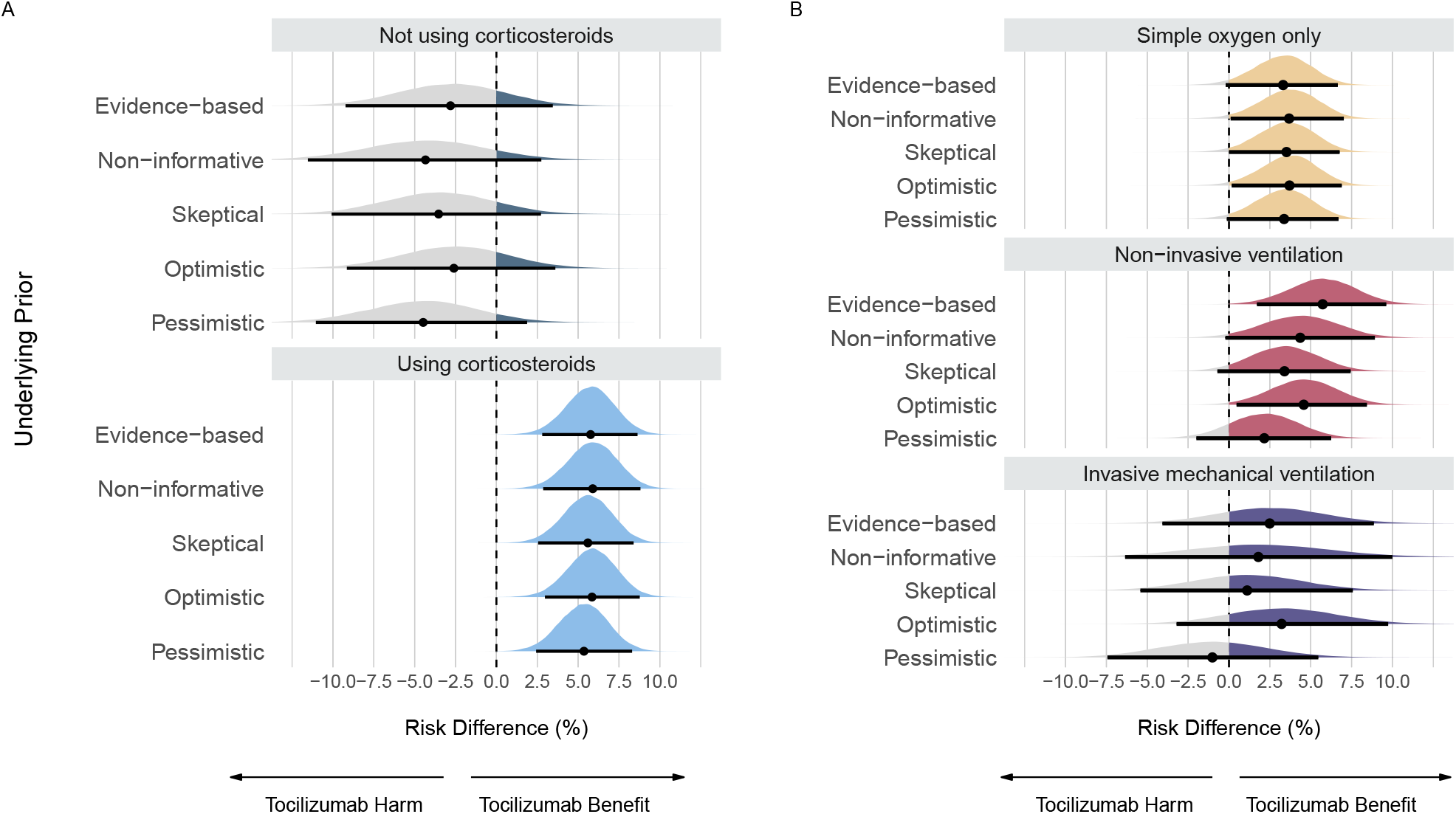
Posterior distributions from sensitivity analyses using different priors on the mortality outcome. Panel A shows posterior distributions on subgroups regarding use of corticosteroids. Panel B shows posterior distributions on subgroups regarding respiratory support. Point estimates depict the median and interval bars depict the 95% highest density intervals.

Figure 4 shows the posterior probabilities for the different subgroups as a function of sensitivity analyses with different baseline risks. The posterior probabilities for any benefit (risk difference ≥ 0%) did not change regardless of the baseline risk in all subgroups. Patients using corticosteroids (Figure 4B) or on non-invasive ventilation (Figure 4D) were particularly insensitive to baseline risks with posterior probabilities of >90% for benefits as large as 2%, provided the baseline risk remained above 15% and 22%, respectively.

**Figure 4.**
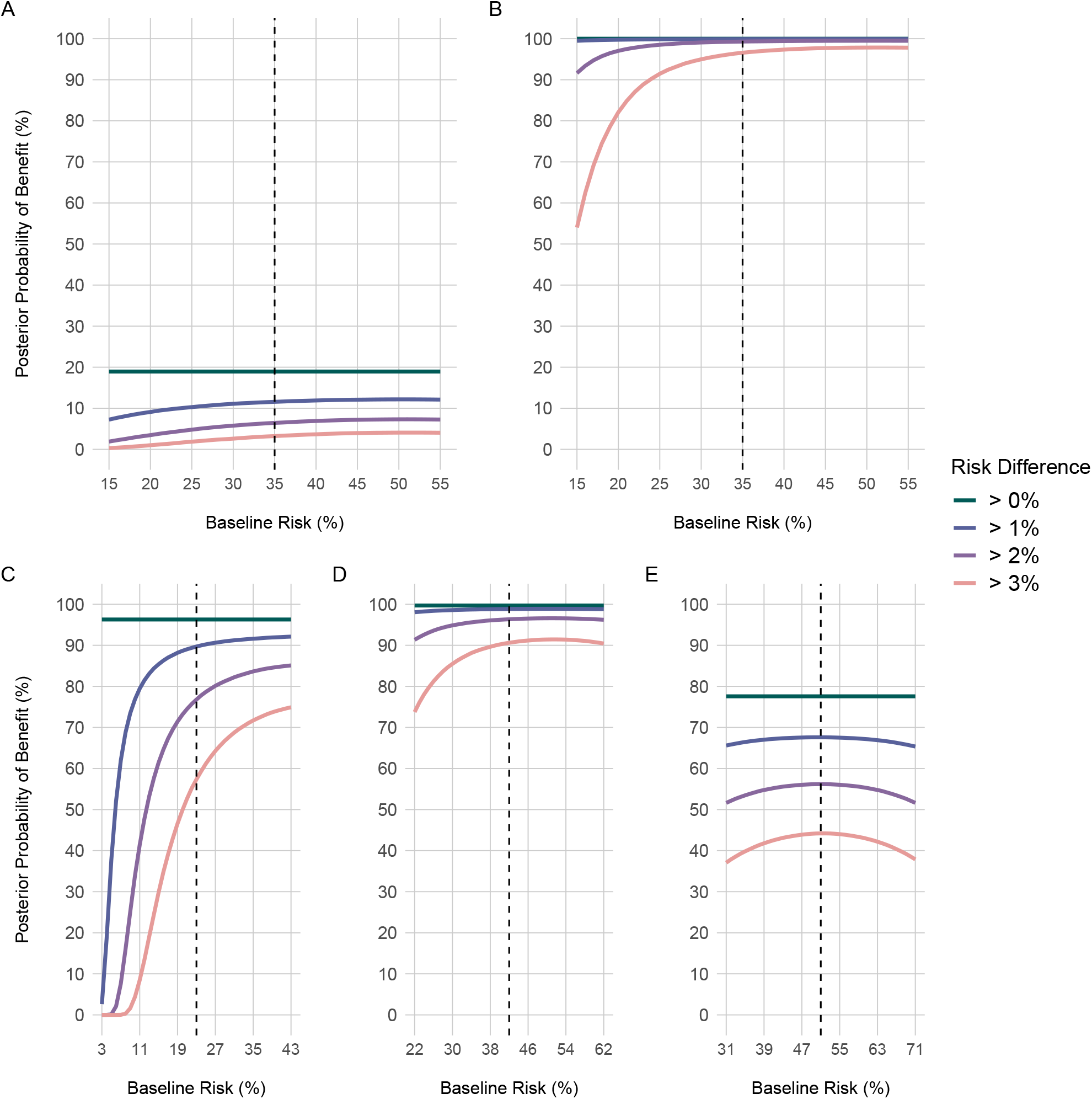
Posterior probabilities from sensitivity analyses using multiple different baseline risks on the mortality outcome. Panel A shows results for patients not using corticosteroids; Panel B shows results for patients using corticosteroids. Panel C shows results for simple oxygen only; Panel D shows results for non-invasive ventilation; Panel E shows results for invasive mechanical ventilation. Each line represents the posterior probability of benefit for a specific cutoff, such as risk difference lower to 0%, 1%, 2% and 3%. Vertical black dashed lines represent the respective baseline risk underlying other analyses (Figure 2), i.e., risk in the control group in the RECOVERY trial for each subgroup (Table 1).

### Hospital discharge

Overall, there was > 95% posterior probabilities of > 2% absolute difference in earlier hospital discharge for patients receiving corticosteroids and for those receiving simple oxygen or non-invasive ventilation therapy. These results were robust while using different prior distributions. Extensive details can be found in the Supplementary Material (Supplementary Tables 5 – 8 and Figures 3 – 8).

## Discussion

Herein we report our Bayesian reanalysis of COVID-19 subgroups from the RECOVERY tocilizumab trial using a variety of transparent priors. For the mortality outcome, the posterior probabilities of any benefit (risk difference ≤ 0%) along with evidence-based priors were strikingly different between patients using and not using corticosteroids (>99% versus 19%, respectively). While the point estimates for benefit with tocilizumab were similar across different respiratory risk profiles, the probabilities of clinically meaningful effects varied substantially, perhaps in part due to differences in sample size and statistical power. Patients on non-invasive ventilation showed the largest benefit with tocilizumab and the most certainty of being associated with a clinically meaningful effect, indicated by 90% probability that the risk difference is greater than 3% (NNT ≤ 33) with the evidence-based prior. Patients receiving simple oxygen, also showed a high (>95%) probability of any benefit with tocilizumab but less certainty that the benefit was as large as NNT of 33 (< 60%). The least certainty of tocilizumab benefit was seen in the group receiving invasive mechanical ventilation where there was a 77% probability of any benefit but a less than 45% probability that its magnitude was as great as a NNT of 33. Lastly, sensitivity analyses showed that the probabilities of any benefit are consistent between priors and baseline risks across most subgroups. In particular, these analyses highlighted the uncertainty around data on invasive mechanical ventilation and the lack of conclusive evidence in favor of tocilizumab’s benefit in this subgroup.

### Comparison with other studies

In the RECOVERY publication,[1] the authors state for tocilizumab’s effect for mortality, “We observed similar results across all prespecified subgroups … Given the number of hypothesis tests done, the suggestion of a larger proportional mortality reduction among those receiving a corticosteroid compared with those not (interaction p=0·01) might reflect the play of chance.” Here, we present a more in-depth analysis and interpretation of these results, casting doubt about the claim of tocilizumab’s universal benefit across sub-groups. First, we show that posterior probabilities of small and large benefits are robustly different between patients using or not corticosteroids. Second, although point estimates are similar between respiratory support subgroups, the posterior probabilities of larger effect sizes are notably distinct, contradicting RECOVERY’s general conclusion applied to all subgroups. Ultimately, we revealed a clear need for more evidence of tocilizumab’s benefit in patients on invasive mechanical ventilation.

A meta-analysis on COVID-19 evaluated the interaction between interleukin-6 inhibitors, combining both tocilizumab and sarilumab, with disease severity and concomitant corticosteroid use on mortality.[26] By applying a Bayesian hierarchical meta-regression model, they have found that “there was no difference” for these subgroups in the odds ratio scale. On the other hand, we report strikingly different posterior distributions of tocilizumab’s effect between the patients using or not corticosteroids and on distinct respiratory support (Figure 1). There may be several explanations for this difference in results. First, the previous meta-analysis included a study on sarilumab with 836 patients, mixing different drugs to the same analysis. Second, our analyses included three additional trials.[16, 20, 24] Third and most importantly, different statistical models were used. While they evaluated the interaction between treatment and subgroup class, we separately estimated the posterior distributions for each subgroup. Moreover, we also transformed results to the risk difference scale and performed multiple sensitivity analyses. In summary, our study provides more easily and clinically interpretable results than previous analyses and underscores how tocilizumab can differently benefit subgroups of patients.

Previous research on COVID-19 has shown that corticosteroids, when not combined with tocilizumab, also reduce deaths, especially in patients with more severe disease.[27] These results could be explained by the direct association of COVID-19’s disease severity with the degree of inflammation. Paradoxically, our results suggest that tocilizumab, also an anti-inflammatory drug, reduces deaths with more certainty and larger effect size in patients with less severe disease than in critically ill patients on invasive mechanical ventilation. Moreover, the low probability of tocilizumab’s benefit in patients not using corticosteroids (Supplementary Table 4) highlights the essential role of the latter in the treatment of COVID-19.[15] Thus, our results reinforce previous evidence on corticosteroids’ efficacy and raise questions about the relationship of anti-inflammatory drugs with the pathophysiology of COVID-19.

In addition to reducing deaths, our study suggests that tocilizumab increases hospital discharge rate. Another meta-analysis has found that tocilizumab has little or no benefit in clinical improvement when analyzing pooled results, defined by proxy outcomes such as hospital discharge.[11] In contrast to these results, we found that the posterior probabilities of benefit in clinical improvement are >90% in patients on different respiratory support. There are also >90% posterior probabilities of benefit as large as NNT of 33 in patients on simple oxygen only or on non-invasive ventilation using evidence-based priors (Supplementary Figure 4D). Our analyses show that these posterior probabilities are stable to the choice of prior, indicating the robustness of our results (Supplementary Table 8). Thus, tocilizumab most likely provides a relevant clinical improvement in patients at different clinical severity, except for the invasive ventilation group and the non-corticosteroid users where additional data are needed.

### Strengths and limitations

Our study has several strengths. Most importantly, Bayesian analyses permit the inclusion of prior beliefs and or prior objective evidence. We only included data from RCTs with a low or moderate risk of bias, as assessed by the living systematic review from which we based our data extraction process.[11] Further, we tried to mitigate potential biases associated with researcher degrees of freedom by pre-registering the data extraction process and part of the analysis plan,[14] and restricting our analyses to RECOVERY specified subgroups. Next, in contrast to P-values and confidence intervals, Bayesian analyses can provide intuitive and understandable probability estimates of each subgroup’s treatment effect. Moreover, these analyses not only inform the probability of any benefit but also calculation of the probabilities of clinically meaningful benefits. This enhances the readers’ ability to distinguish clinical from statistical significance. Lastly, we incorporated most of the high-quality evidence available on tocilizumab and COVID-19 to our prior distributions.

Notwithstanding these strengths, our study also has limitations. First, we did not have access to patient-level data for any RCTs included in this article. Subgroup analyses that separate patients by a single baseline characteristic are over-simplified and can bring shortcomings.[2] Lack of patient-level data does not allow analyses using more complex statistical models that incorporate multiple characteristics.[28] Second, our Bayesian analyses were not planned when the RECOVERY was published and thus must be interpreted as exploratory. Third, is the classical argument of Bayesian subjectivity due to the need for priors.[29] Although we agree with need for transparency and caution in the choice of priors, the synthesis of objective prior evidence with current data is likely to furnish the most accurate and least biased treatment estimates. Bayesian analyses are not necessarily more subjective than frequentist methods, simply more explicit about the underlying assumptions. By using multiple priors, incorporating data from previously published RCTs, following recent recommendations for Bayesian analyses,[13] and explicitly stating how our priors are defined, we argue that Bayesian analyses are, in fact, more transparent and robust than usual analyses. Fourth, we limited our focus to benefit-related outcomes and did not analyze results regarding adverse effects. Fifth, because there was one study with high-risk of bias or we could not extract subgroup-specific data, we did not include a total of four studies that contributed to 46% of weight in the meta-analysis, used as our reference,[11] regarding hospital discharge.[19, 22, 23, 25] Although we could not also include six studies with all-cause mortality data, these trials only contributed to less than 7% of weight in the meta-analysis (Supplementary Material).[18, 19, 21–23, 25]

## Conclusions

In conclusion, Bayesian methods overcome the standard null hypothesis significance testing limitations by estimating the probability of benefit and harm while incorporating prior evidence. By highlighting the difference between statistically and clinically significant results, Bayesian analyses provide additional insights on traditional RCT subgroup analyses. Specifically, COVID-19 hospitalized patients exposed to corticosteroids have a high probability of a clinically meaningful mortality benefit from tocilizumab. Similarly, tocilizumab offers a high probability of a clinically significant mortality reduction in patients on simple oxygen only or non-invasive ventilation, especially for the latter. Tocilizumab also most likely increases clinical improvement (hospital discharge) in hospitalized COVID-19 patients. Future research should further address if patients on invasive mechanical ventilation can also benefit from tocilizumab.

## Data Availability

The data and full analysis code are available at https://bit.ly/3xnTJSV.

