## Supplementary Material for "Tocilizumab in COVID-19 – A Bayesian reanalysis of RECOVERY"

**Arthur M. Albuquerque**
School of Medicine, Universidade Federal do Rio de Janeiro, Brazil

**Lucas Tramujas, MD**
HCor Research Institute, São Paulo, Brazil

**Lorenzo R. Sewanan, MD, PhD**
Department of Internal Medicine, Columbia University, U.S.A.

**James M. Brophy, MD, PhD**
McGill University Health Center, Canada


### Summary

**Data extraction** 2

**Overall characteristics** 10

**Statistical analyses** 13

**Results: Mortality outcome** 15

#### Data extraction

In this section, we discuss about our data extraction process.

In brief, we pre-registered our protocol in the OpenScienceFramework (https://osf.io/cytq8/) before any analyses were performed. We extracted summary data from randomized controlled trials (RCT) included in a living systematic review on Cochrane. We accessed each individual RCT manuscript to extract data. We contacted the authors by e-mail to obtain relevant data not available in the manuscript. However, we did not have success to obtain any extra information by e-mail.

In the following pages, we further describe our data extraction process. First, we reproduce the living systematic review (meta-analyses) from which we extracted the data for mortality ("All-cause mortality D28") and hospital discharge ("Clinical Improvement D28"). We then summarize our workflow on article screening and data extraction with a PRISMA diagram. Next, we show tables in which we discuss details on studies that were included or excluded from our analyses. The "Source" columns regard the figure or table from which we extracted summary data of each manuscript. Lastly, we elaborate the details about the data extraction and analysis of a specific RCT.

Data: Thu Van Nguyen, Gabriel Ferrand, Sarah Cohen-Boulakia, Ruben Martinez, Philipp Kapp, Emmanuel Coquery, ... for the COVID-NMA consortium. (2020). RCT studies on preventive measures and treatments for COVID-19 [Data set]. Zenodo. http://doi.org/10.5281/zenodo.4266528 ; Visualizations: Romain Vuillemot - LIRIS, École Centrale de Lyon; Philippe Rivière - LIRIS, VisionsCarto; Pierre Ripoll - LIRIS, INSA Lyon; Julien Barnier - Centre Max Weber, CNRS. Retrieved from: 'https://covid-nma.com/dataviz/' [Online Resource]

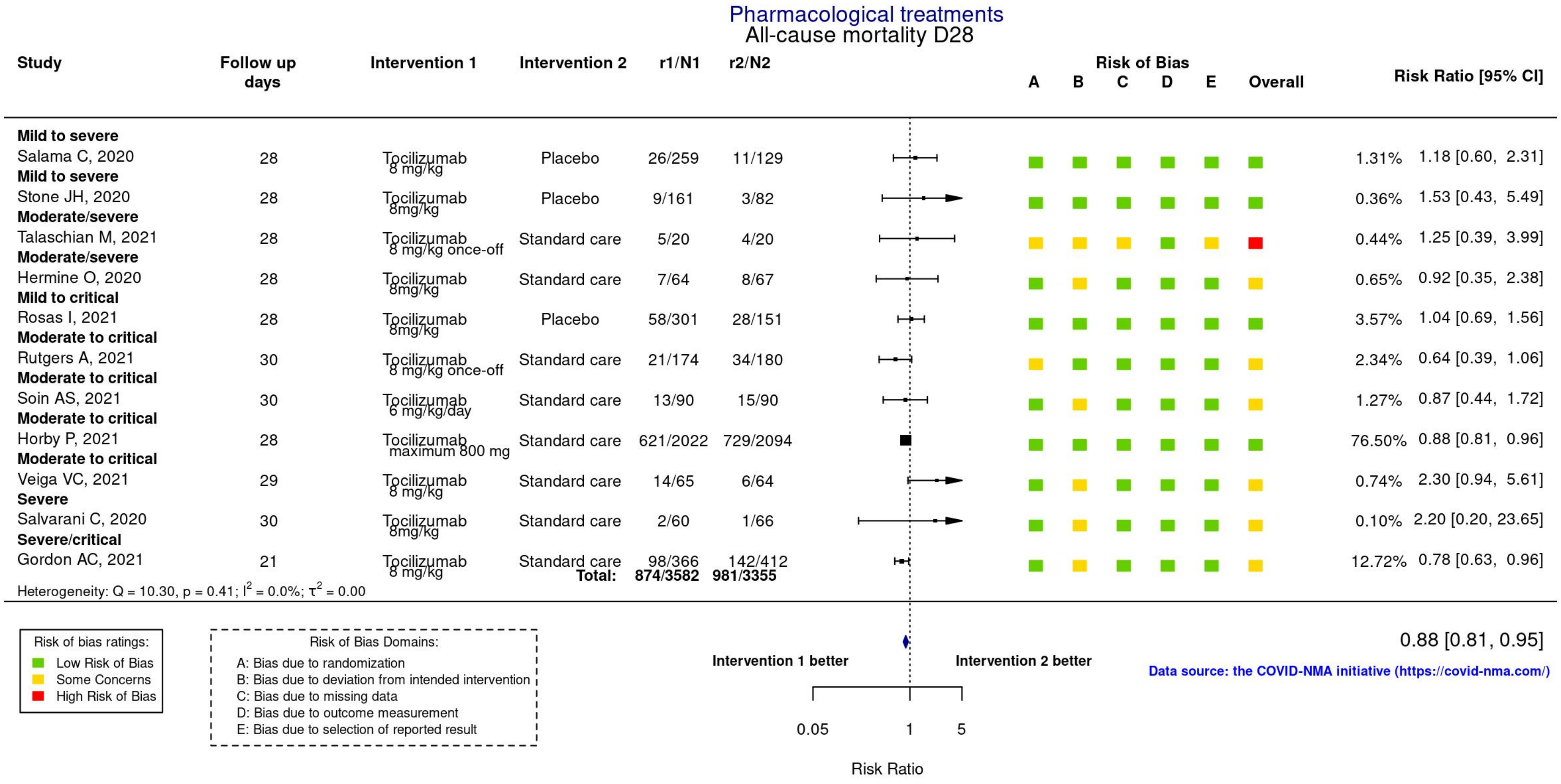

Data: Thu Van Nguyen, Gabriel Ferrand, Sarah Cohen-Boulakia, Ruben Martinez, Philipp Kapp, Emmanuel Coquery, ... for the COVID-NMA consortium. (2020). RCT studies on preventive measures and treatments for COVID-19 [Data set]. Zenodo. http://doi.org/10.5281/zenodo.4266528 ; Visualizations: Romain Vuillemot - LIRIS, École Centrale de Lyon; Philippe Rivière - LIRIS, VisionsCarto; Pierre Ripoll - LIRIS, INSA Lyon; Julien Barnier - Centre Max Weber, CNRS. Retrieved from: 'https://covid-nma.com/dataviz/' [Online Resource]

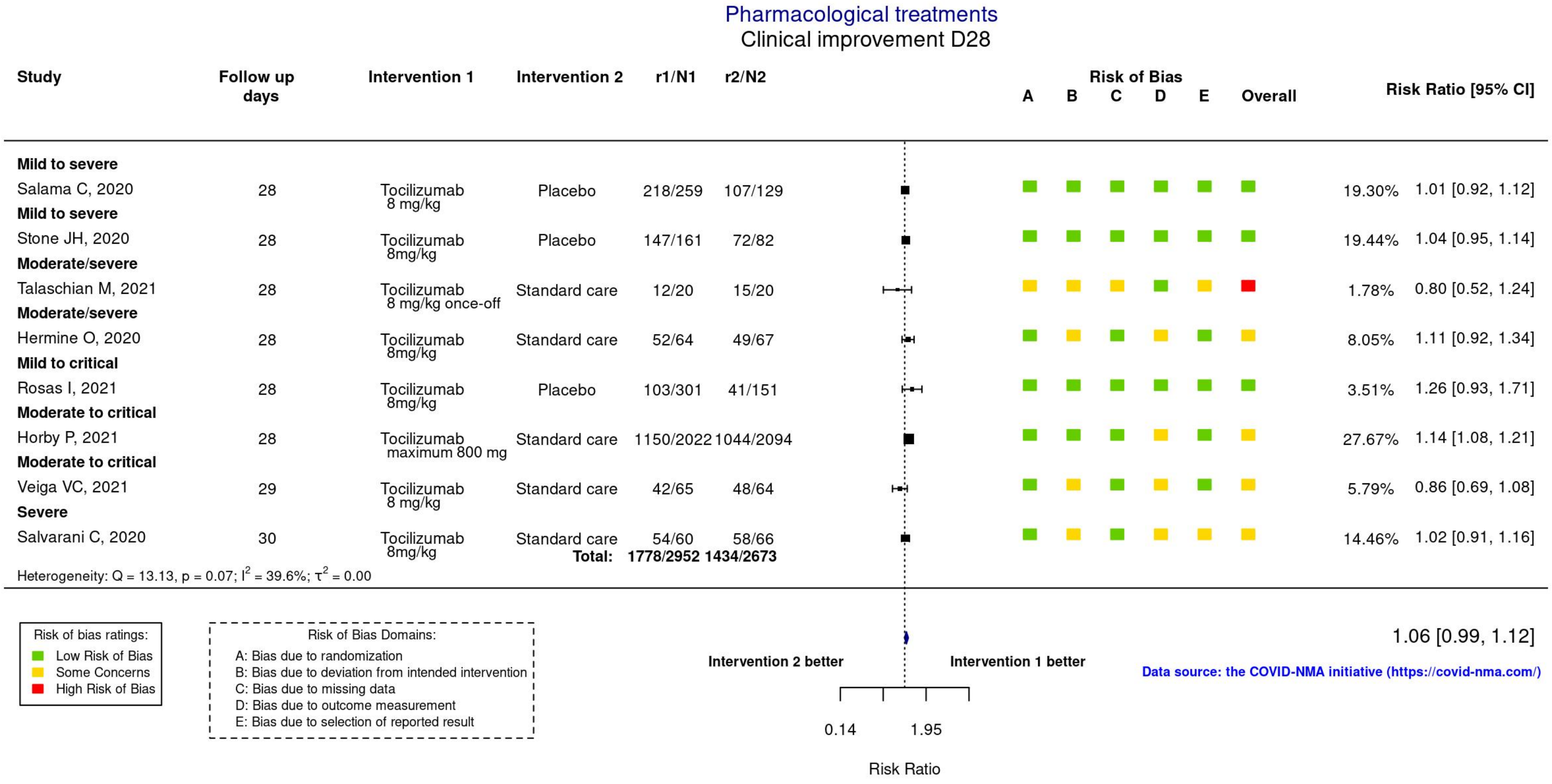

The diagram below show the workflow regarding total studies that were used to create prior distributions regardless of the outcome.

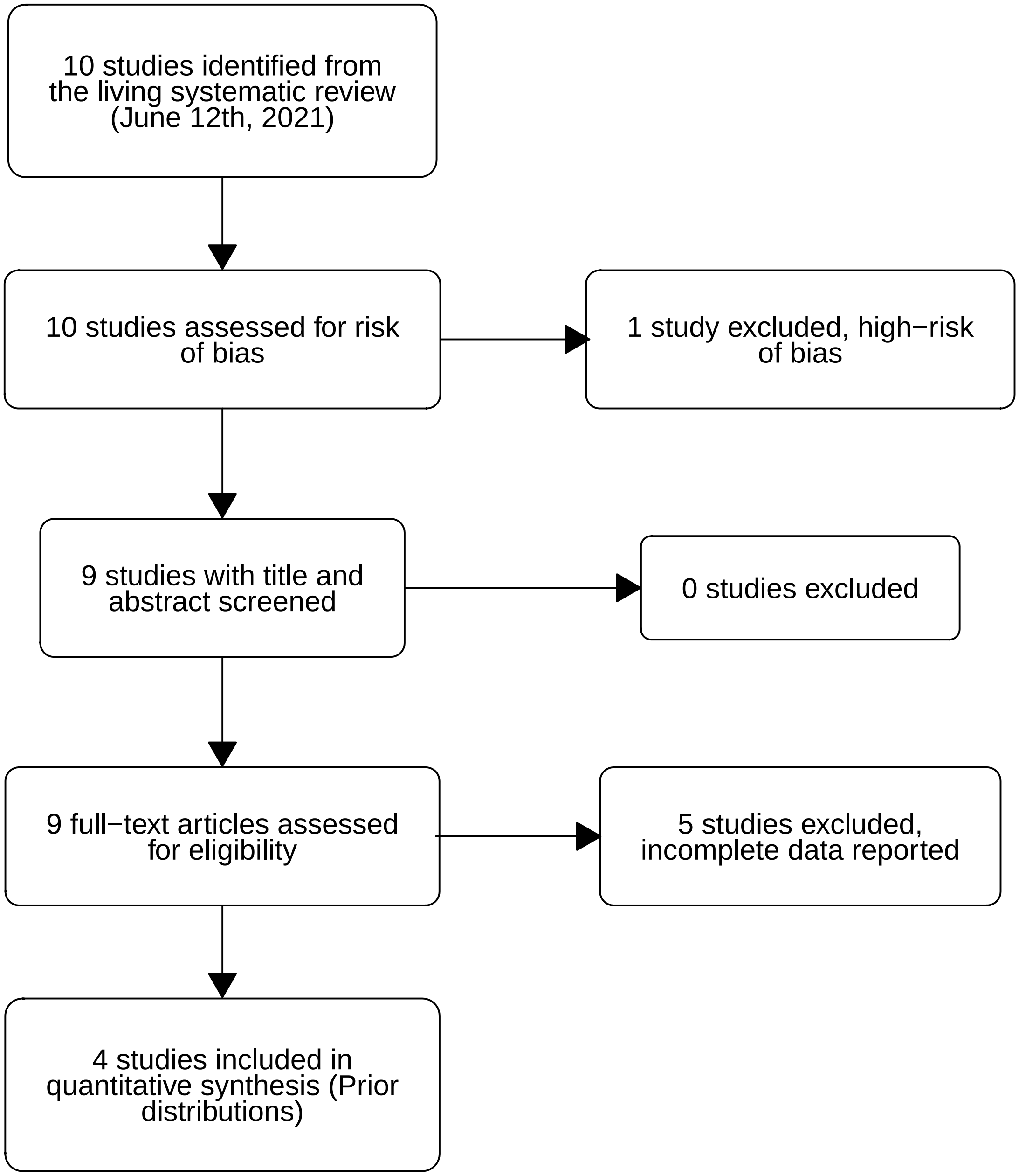

#### Included studies for the mortality outcome

| Study | Source | Notes |
| --- | --- | --- |
| **Not using corticosteroids** | | |
| RECOVERY | Figure 3 | “Use of corticosteroids” section |
| COVACTA | Table S10 | Ordinal category 7 means death. |
| REMAP-CAP | Figure 9 | Full explanation on the following pages of this supplementary material |
| **Using corticosteroids** | | |
| RECOVERY | Figure 3 | “Use of corticosteroids” section |
| COVACTA | Table S10 | Ordinal category 7 means death. |
| REMAP-CAP | Figure 9 | Full explanation on the following pages of this supplementary material |
| **Simple oxygen only** | | |
| RECOVERY | Figure 3 | “Respiratory support at randomisation” section |
| COVACTA | Figure 2 | Ordinal category 3 means “non–ICU hospitalization with supplemental oxygen”. Thus, Category 3 at baseline equals to the “simple oxygen only” subgroup. Ordinal Category 7 means death. |
| CORIMUNO-19 | Table 2 | This study only included patients “...receiving at least 3L/min oxygen (O2) but without high-flow oxygen (HFO)...”. Thus, we included all data available. |
| **Non-invasive ventilation** | | |
| RECOVERY | Figure 3 | “Respiratory support at randomisation” section |
| COVACTA | Figure 2 | Ordinal category 4 means “ICU or non–ICU hospitalization with noninvasive ventilation or high-flow oxygen”. Thus, Category 4 at baseline equals to the “non-invasive ventilation” subgroup. Ordinal Category 7 means death. |
| REMAP-CAP | Table S7 | “Progression to invasive mechanical ventilation, ECMO or death, restricted to those not intubated at baseline” section |
| Salvarani | Table 2 | This study only included patients “Patients at enrollment were allowed to receive oxygen therapy with Venturi mask or high-flow nasal cannula with recorded and preset FIO2...”. Thus, we included all data available. |
| **Invasive mechanical ventilation** | | |
| RECOVERY | Figure 3 | “Respiratory support at randomisation” section |
| COVACTA | Figure 2 | Ordinal category 5 means “ICU hospitalization with mechanical ventilation” and category 6 means “ICU hospitalization with extracorporeal membrane oxygenation or mechanical ventilation and additional organ support”. Thus, both categories combined equals to the “invasive mechanical ventilation” subgroup. Ordinal Category 7 means death. |
| REMAP-CAP | Table 2 and Table S7 | Table S7 shows how many patients that were free of IMV at baseline died (53 and 82) and their sample size (242 + 273). Table 2 shows total number of deaths (98 and 142) and total sample size (350 + 397). Thus, we subtracted values between tables to get how many patients on IMV at baseline died and sample size. |

### Included studies for the hospital discharge outcome

| Study | Source | Notes |
|---|---|---|
| **Not using corticosteroids** | | |
| RECOVERY | Webfigure 1 | “Use of corticosteroids” section |
| COVACTA | Table S10 | Ordinal category 1 means “discharged or ready for discharge” |
| **Using corticosteroids** | | |
| RECOVERY | Webfigure 1 | “Use of corticosteroids” section |
| COVACTA | Table S10 | Ordinal category 1 means “discharged or ready for discharge” |
| **Simple oxygen only** | | |
| RECOVERY | Webfigure 1 | “Respiratory support at randomisation” section |
| COVACTA | Figure 2 | Ordinal category 3 means “non–ICU hospitalization with supplemental oxygen”. Thus, Category 3 at baseline equals to the “simple oxygen only” subgroup. Ordinal Category 7 means “discharged or ready for discharge”. |
| CORIMUNO-19 | eTable 9 | This study only included patients “...receiving at least 3L/min oxygen (O2) but without high-flow oxygen (HFO)...”. Thus, we included all data available. |
| **Non-invasive ventilation** | | |
| RECOVERY | Webfigure 1 | “Respiratory support at randomisation” section |
| COVACTA | Figure 2 | Ordinal category 4 means “ICU or non–ICU hospitalization with noninvasive ventilation or high-flow oxygen”. Thus, Category 4 at baseline equals to the “non-invasive ventilation” subgroup. Ordinal Category 7 means “discharged or ready for discharge”. |
| Salvarani | Table 2 | This study only included patients “Patients at enrollment were allowed to receive oxygen therapy with Venturi mask or high-flow nasal cannula with recorded and preset FIO2...”. Thus, we included all data available. |
| **Invasive mechanical ventilation** | | |
| RECOVERY | Webfigure 1 | “Respiratory support at randomisation” section |
| COVACTA | Figure 2 | Ordinal category 5 means “ICU hospitalization with mechanical ventilation” and category 6 means “ICU hospitalization with extracorporeal membrane oxygenation or mechanical ventilation and additional organ support”. Thus, both categories combined equals to the “invasive mechanical ventilation” subgroup. Ordinal Category 7 means “discharged or ready for discharge”. |

Excluded studies for the mortality outcome

| Study | Notes |
|---|---|
| **Use of corticosteroids subgroups** | |
| TOCIBRAS | Not possible to extract data per subgroup |
| Stone | Not possible to extract data per subgroup |
| EMPACTA | Not possible to extract data per subgroup |
| COVINTOC | Not possible to extract data per subgroup |
| Rutgers | Not possible to extract data per subgroup |
| **Respiratory support subgroups** | |
| TOCIBRAS | Not possible to extract data per subgroup |
| Stone | Not possible to extract data per subgroup |
| EMPACTA | Not possible to extract data per subgroup |
| COVINTOC | Not possible to extract data per subgroup |
| Rutgers | Not possible to extract data per subgroup |

Excluded studies for the hospital discharge outcome

| Study | Notes |
|---|---|
| **Use of corticosteroids subgroups** | |
| REMAP-CAP | Not possible to extract data per subgroup |
| TOCIBRAS | Not possible to extract data per subgroup |
| Stone | Not possible to extract data per subgroup |
| EMPACTA | Not possible to extract data per subgroup |
| **Respiratory support subgroups** | |
| REMAP-CAP | Not possible to extract data per subgroup |
| TOCIBRAS | Not possible to extract data per subgroup |
| Stone | Not possible to extract data per subgroup |
| EMPACTA | Not possible to extract data per subgroup |

As mentioned in one of the tables above, we will now discuss further details about data on the use of corticosteroids subgroups from REMAP-CAP.

The relevant data from these subgroups are shown in Figure 9 in REMAP-CAP's supplementary material. Unfortunately, the authors do not provide the raw number of events per subgroup. Instead, the data is displayed visually in stacked bars. Thus, we used the {juicr} package to extract data directly from Figure 9. The raw data and extraction report generated by {juicr} can be found in this project's GitHub repository.

Of note, REMAP-CAP was a three-arm randomized controlled trial that tested tocilizumab and sarilumab (both are IL-6 antagonists) vs. usual care. The authors only provided pooled data from these IL-6 antagonists vs. usual care in Figure 9. Thus, we decided to perform sensitivity analyses with this data to dampen the influence of sarilumab in final results.

We created evidence-based priors for these subgroups using data from REMAP-CAP with different weights: 100%, 75% and 50%. This approach implies that the numbers of events and sample sizes from REMAP-CAP were multiplied by 1, 0.75 or 0.5, respectively.

As mentioned in the Methods section of this article, we pooled data from similar studies with a random-effect meta-analysis using a restricted maximum-likelihood estimator to create evidence-based priors. We will now display evidence-based prior distributions for both "Not using corticosteroids" and "Using corticosteroids" subgroups that were generated with different weights on REMAP-CAP. Top panels regards "Not using corticosteroids" and the bottom panels "Using corticosteroids". We show these distributions in both log-odds ratio and odds ratio scales to ease interpretation.

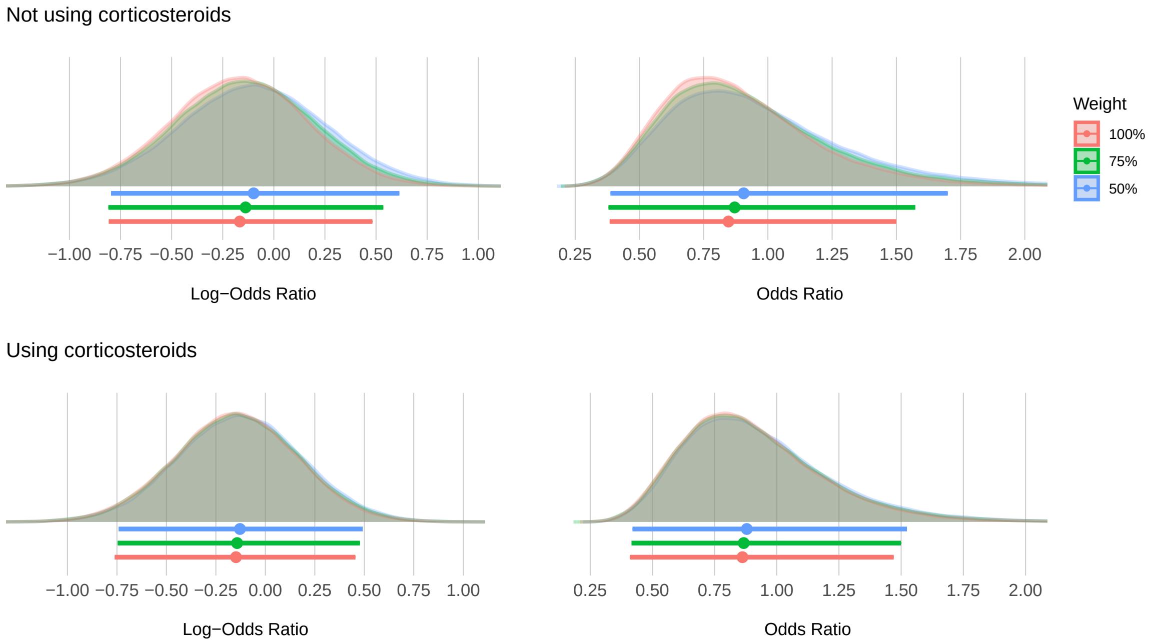

Point estimates depict the median and interval bars depict the 95% highest density interval.

Because prior distributions are notably similar regardless of the weight used, we have decided to only use the ones with 100% weight for our analyses.

#### Overall characteristics

In this section, we show study and patient characteristics regarding the data used for prior distribution elicitation.

Included studies

| Study Characteristic | |
|---|---|
| **RECOVERY** | |
| Year | 2021 |
| Trial design | Open-label |
| Follow-up period (days) | 28 |
| Control treatment | Standard of care |
| **COVACTA** | |
| Year | 2021 |
| Trial design | Double-blinded |
| Follow-up period (days) | 28 |
| Control treatment | Placebo |
| **REMAP-CAP** | |
| Year | 2021 |
| Trial design | Open-label |
| Follow-up period (days) | 21 |
| Control treatment | Standard of care |
| **CORIMUNO-19** | |
| Year | 2020 |
| Trial design | Open-label |
| Follow-up period (days) | 28 |
| Control treatment | Standard of care |
| **Salvarani** | |
| Year | 2020 |
| Trial design | Open-label |
| Follow-up period (days) | 30 |
| Control treatment | Standard of care |

#### Patient characteristics of included studies

(Filled circles indicate what subgroups from each trial were used in our analyses)

| | Interventions | | Outcomes | |
|---|---|---|---|---|
| Study Characteristic | Control | Tocilizumab | Mortality | Hospital Discharge |
| **RECOVERY** | | | | |
| Number of patients | 2094 | 2022 | | |
| Age (SD) | 64 (14) | 63 (14) | | |
| Male sex (%) | 69 | 66 | | |
| Confirmed SARS-CoV-2 infection (%) | 96 | 95 | | |
| Use of corticosteroids (%) | 82 | 82 | ● | ● |
| Simple oxygen only at randomisation (%) | 45 | 46 | ● | ● |
| Non-invasive ventilation at randomisation (%) | 41 | 41 | ● | ● |
| Invasive mechanical ventilation at randomisation (%) | 14 | 13 | ● | ● |
| **COVACTA** | | | | |
| Number of patients | 144 | 294 | | |
| Age (SD) | 61 (14) | 60 (15) | | |
| Male sex (%) | 70 | 70 | | |
| Confirmed SARS-CoV-2 infection (%) | 100 | 100 | | |
| Use of corticosteroids (%) | 28.5 | 19 | ● | ● |
| Simple oxygen only at randomisation (%) | 31 | 26.5 | ● | ● |
| Non-invasive ventilation at randomisation (%) | 27 | 32 | ● | ● |
| Invasive mechanical ventilation at randomisation (%) | 38 | 38 | ● | ● |
| **REMAP-CAP** | | | | |
| Number of patients | 402 | 353 | | |
| Age (SD) | 61 (13) | 61 (12) | | |
| Male sex (%) | 70 | 74 | | |
| Confirmed SARS-CoV-2 infection (%) | 85 | 82 | | |
| Use of corticosteroids (%) | $> 80$ | $> 80$ | ● | ○ |
| Simple oxygen only at randomisation (%) | $< 1$ | $< 1$ | ○ | ○ |
| Non-invasive ventilation at randomisation (%) | 69 | 71 | ● | ○ |
| Invasive mechanical ventilation at randomisation (%) | 30 | 29 | ● | ○ |
| **CORIMUNO** | | | | |
| Number of patients | 67 | 63 | | |
| Age (IQR) | 63 (57 - 72) | 64 (57 - 74) | | |
| Male sex (%) | 66 | 70 | | |
| Confirmed SARS-CoV-2 infection (%) | 90 | 89 | | |
| Use of corticosteroids (%) | 61 | 33 | ○ | ○ |
| Simple oxygen only at randomisation (%) | 100 | 100 | ● | ● |
| Non-invasive ventilation at randomisation (%) | 0 | 0 | ○ | ○ |
| Invasive mechanical ventilation at randomisation (%) | 0 | 0 | ○ | ○ |
| **Salvarani** | | | | |
| Number of patients | 62 | 60 | | |
| Age (IQR) | 60 (54 - 69) | 61 (51 - 73) | | |
| Male sex (%) | 56 | 67 | | |
| Confirmed SARS-CoV-2 infection (%) | 100 | 100 | | |
| Use of corticosteroids (%) | 11 | 10 | ○ | ○ |
| Simple oxygen only at randomisation (%) | 0 | 0 | ○ | ○ |
| Non-invasive ventilation at randomisation (%) | 100 | 100 | ● | ● |
| Invasive mechanical ventilation at randomisation (%) | 0 | 0 | ○ | ○ |

#### Statistical analyses

In this section, we will briefly explain our statistical analyses. First, we pooled data from similar studies with a random-effect meta-analysis using a restricted maximum-likelihood estimator to create a evidence-based prior for each subgroup, as depicted below:

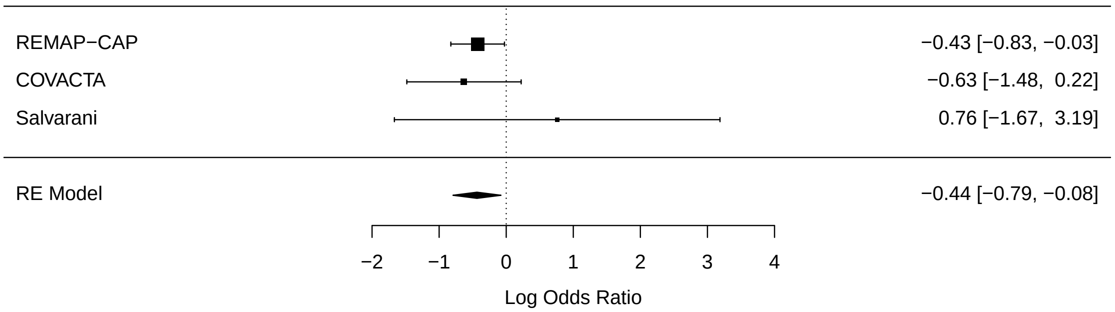

In this figure, we show only one prior distribution as an example. Random-effects (RE) Model represents the evidence-based prior distribution for the non-invasive ventilation subgroup on the mortality outcome.

Next, we calculated the log-odds ratio for each subgroup in RECOVERY:

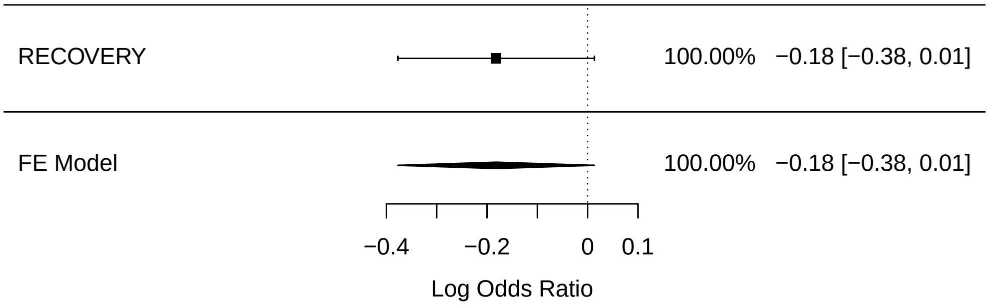

In this figure, we show only one likelihood distribution as an example. Fixed-effects (FE) Model represents the likelihood distribution for the non-invasive ventilation subgroup on the mortality outcome.

We then estimated the posterior distribution for each subgroup by calculating its mean and variance. Following Spiegelhalter et al., we assumed that every prior and likelihood distribution (in the log-odds ratio scale) to be normal distributed. Thus, each prior distribution can be described as $N(\theta, \sigma^2)$, and each likelihood distribution as $N(\hat{\theta}, \hat{\sigma}^2)$.

Using conjugate normal analysis, we then estimated the mean and variance of each posterior distribution with the following formulas:

- Mean: $\dfrac{\frac{\theta}{\sigma^2} + \frac{\hat{\theta}}{\hat{\sigma}^2}}{\frac{1}{\sigma^2} + \frac{1}{\hat{\sigma}^2}}$ ; Variance: $\dfrac{1}{\frac{1}{\sigma^2} + \frac{1}{\hat{\sigma}^2}}$

Next, we drew and exponentiated 100,000 samples from the posterior distributions to change to an odds ratio scale. Lastly, following Doi et al., risk differences were derived using this formula:

$$RD = \frac{r_0\left(r_0 - 1\right)\left(OR - 1\right)}{r_0 OR - r_0 + 1}$$

where $RD$ is risk difference, $r_0$ is the outcome risk in control arm in RECOVERY and $OR$ is odds ratio. Tables below show the risks in the control arm for each subgroup and outcome:

Mortality outcome

| Subgroup | Risk in Control Arm (%) |
|---|---|
| **Use of corticosteroids** | |
| Not using | 35 |
| Using | 35 |
| **Respiratory support** | |
| Simple oxygen only | 23 |
| Non-invasive ventilation | 42 |
| Invasive mechanical | 51 |

Hospital discharge outcome

| Subgroup | Risk in Control Arm (%) |
|---|---|
| **Use of corticosteroids** | |
| Not using | 46 |
| Using | 51 |
| **Respiratory support** | |
| Simple oxygen only | 68 |
| Non-invasive ventilation | 42 |
| Invasive mechanical | 16 |

As described in the Methods section of this article, we re-estimated the posterior distributions using a plethora of different prior distributions and baseline risks for sensitivity analyses.

### Results: Mortality outcome

#### Prior, RECOVERY, and Posterior distributions

##### Supplementary Table 1

| | 95% Highest Density Interval | | |
|---|---|---|---|
| Distribution | Median | Lower limit | Upper limit |
| Not using corticosteroids | | | |
| Prior | 0.85 | 0.44 | 1.61 |
| RECOVERY | 1.21 | 0.89 | 1.64 |
| Posterior | 1.13 | 0.86 | 1.48 |
| Using corticosteroids | | | |
| Prior | 0.86 | 0.47 | 1.57 |
| RECOVERY | 0.76 | 0.66 | 0.88 |
| Posterior | 0.77 | 0.67 | 0.88 |
| Simple oxygen only | | | |
| Prior | 1.36 | 0.50 | 3.83 |
| RECOVERY | 0.80 | 0.64 | 1.00 |
| Posterior | 0.82 | 0.66 | 1.02 |
| Non-invasive ventilation | | | |
| Prior | 0.65 | 0.45 | 0.92 |
| RECOVERY | 0.83 | 0.68 | 1.01 |
| Posterior | 0.79 | 0.66 | 0.93 |
| Invasive mechanical ventilation | | | |
| Prior | 0.86 | 0.56 | 1.31 |
| RECOVERY | 0.93 | 0.67 | 1.29 |
| Posterior | 0.91 | 0.70 | 1.18 |

95% highest density intervals of evidence-based prior, RECOVERY, and posterior distributions on the mortality outcome. This table complements Figure 1.

#### Posterior probabilities using evidence-based priors

##### Supplementary Table 2

| | 95% Highest Density Interval | | | Probability of Risk Difference ≥ X% | | | |
|---|---|---|---|---|---|---|---|
| Subgroup | Median | Lower limit | Upper limit | Pr(≥ 0%) | Pr(≥ 1%) | Pr(≥ 2%) | Pr(≥ 3%) |
| Use of corticosteroids | | | | | | | |
| Not using | -2.8 | -9.2 | 3.5 | 18.9 | 11.5 | 6.3 | 3.1 |
| Using | 5.8 | 2.8 | 8.6 | 100.0 | 99.9 | 99.3 | 96.5 |
| Respiratory support | | | | | | | |
| Simple oxygen only | 3.3 | -0.2 | 6.7 | 96.2 | 89.7 | 76.6 | 57.0 |
| Non-invasive ventilation | 5.7 | 1.7 | 9.6 | 99.7 | 98.9 | 96.4 | 90.6 |
| Invasive mechanical ventilation | 2.5 | -4.1 | 8.9 | 77.3 | 67.3 | 55.8 | 44.0 |

95% highest density intervals and posterior probabilities of benefit on the mortality outcome. This table complements Figure 2.

### Sensitivity analyses using different priors

#### Supplementary Figure 1

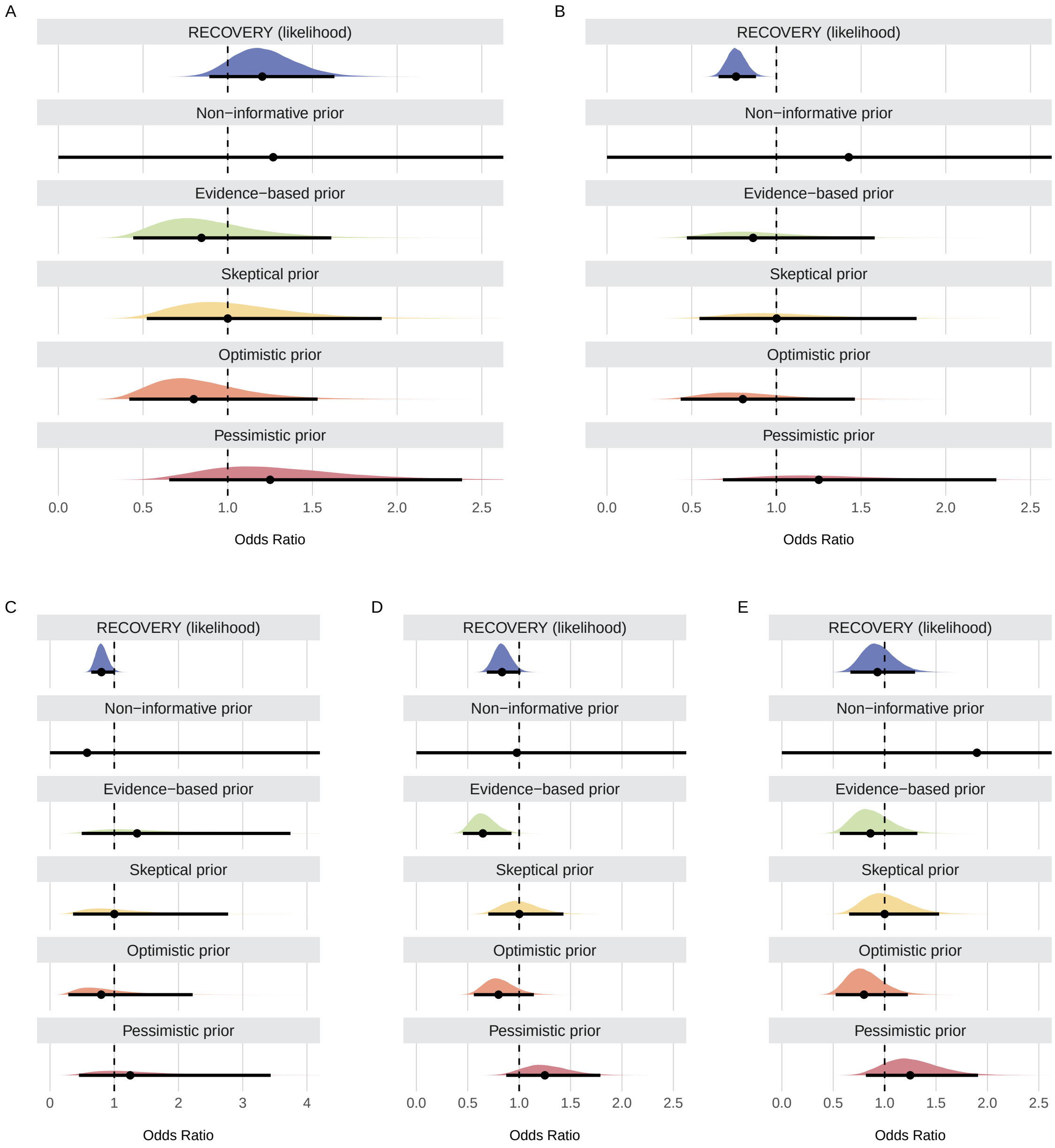

Prior distributions for each subgroup on the mortality outcome. Point estimates depict the median and interval bars depict 95th quantile intervals. Panel A shows results for patients not using corticosteroids; Panel B shows results for patients using corticosteroids. Panel C shows results for simple oxygen only; Panel D shows results for non-invasive ventilation; Panel E shows results for invasive mechanical ventilation.

**Supplementary Table 3**

| Underlying Prior | 95% Highest Density Interval | | |
|---|---|---|---|
| | Median | Lower limit | Upper limit |
| Not using corticosteroids | | | |
| Evidence-based | -2.8 | -9.2 | 3.5 |
| Non-informative | -4.3 | -11.5 | 2.7 |
| Skeptical | -3.5 | -10.1 | 2.7 |
| Optimistic | -2.6 | -9.2 | 3.6 |
| Pessimistic | -4.5 | -11.0 | 1.9 |
| Using corticosteroids | | | |
| Evidence-based | 5.8 | 2.8 | 8.6 |
| Non-informative | 5.9 | 2.9 | 8.8 |
| Skeptical | 5.6 | 2.6 | 8.4 |
| Optimistic | 5.9 | 3.0 | 8.8 |
| Pessimistic | 5.4 | 2.4 | 8.3 |
| Simple oxygen only | | | |
| Evidence-based | 3.3 | -0.2 | 6.7 |
| Non-informative | 3.7 | 0.1 | 7.0 |
| Skeptical | 3.5 | 0.0 | 6.8 |
| Optimistic | 3.7 | 0.2 | 6.9 |
| Pessimistic | 3.4 | -0.1 | 6.7 |
| Non-invasive ventilation | | | |
| Evidence-based | 5.7 | 1.7 | 9.6 |
| Non-informative | 4.4 | -0.2 | 8.9 |
| Skeptical | 3.4 | -0.7 | 7.4 |
| Optimistic | 4.6 | 0.5 | 8.4 |
| Pessimistic | 2.2 | -2.0 | 6.3 |
| Invasive mechanical ventilation | | | |
| Evidence-based | 2.5 | -4.1 | 8.9 |
| Non-informative | 1.8 | -6.4 | 10.0 |
| Skeptical | 1.1 | -5.4 | 7.6 |
| Optimistic | 3.2 | -3.2 | 9.7 |
| Pessimistic | -1.0 | -7.4 | 5.5 |

Median and 95% highest density intervals of posteriors distributions from sensitivity analyses using different priors on the mortality outcome. This table complements Figure 3.

**Supplementary Table 4**

| Underlying Prior | Propability of Harm | | | Probability of Benefit | | | |
|---|---|---|---|---|---|---|---|
| | Pr(< -3%) | Pr(< -2%) | Pr(< -1%) | Pr(> 0%) | Pr(> 1%) | Pr(> 2%) | Pr(> 3%) |
| Not using corticosteroids | | | | | | | |
| Evidence-based | 47.8 | 59.9 | 71.4 | 18.9 | 11.5 | 6.3 | 3.1 |
| Non-informative | 64.3 | 74.2 | 82.3 | 11.4 | 6.8 | 3.7 | 1.9 |
| Skeptical | 56.6 | 68.0 | 78.3 | 13.6 | 7.9 | 4.0 | 1.8 |
| Optimistic | 45.3 | 57.4 | 69.0 | 20.8 | 12.9 | 7.3 | 3.8 |
| Pessimistic | 67.4 | 77.6 | 85.8 | 8.3 | 4.4 | 2.1 | 0.9 |
| Using corticosteroids | | | | | | | |
| Evidence-based | 0.0 | 0.0 | 0.0 | 100.0 | 99.9 | 99.3 | 96.5 |
| Non-informative | 0.0 | 0.0 | 0.0 | 100.0 | 99.9 | 99.3 | 96.8 |
| Skeptical | 0.0 | 0.0 | 0.0 | 100.0 | 99.9 | 99.0 | 95.6 |
| Optimistic | 0.0 | 0.0 | 0.0 | 100.0 | 99.9 | 99.4 | 97.0 |
| Pessimistic | 0.0 | 0.0 | 0.0 | 100.0 | 99.8 | 98.6 | 93.9 |
| Simple oxygen only | | | | | | | |
| Evidence-based | 0.1 | 0.3 | 1.1 | 96.2 | 89.7 | 76.6 | 57.0 |
| Non-informative | 0.1 | 0.2 | 0.8 | 97.4 | 92.6 | 82.2 | 64.8 |
| Skeptical | 0.0 | 0.2 | 0.8 | 97.2 | 91.9 | 80.3 | 61.7 |
| Optimistic | 0.0 | 0.1 | 0.6 | 97.8 | 93.2 | 83.0 | 65.5 |
| Pessimistic | 0.1 | 0.2 | 1.0 | 96.6 | 90.4 | 77.7 | 58.4 |
| Non-invasive ventilation | | | | | | | |
| Evidence-based | 0.0 | 0.0 | 0.1 | 99.7 | 98.9 | 96.4 | 90.6 |
| Non-informative | 0.1 | 0.4 | 1.3 | 96.7 | 92.1 | 84.0 | 71.6 |
| Skeptical | 0.1 | 0.6 | 1.9 | 94.6 | 87.3 | 74.7 | 57.6 |
| Optimistic | 0.0 | 0.1 | 0.4 | 98.6 | 95.7 | 89.3 | 77.7 |
| Pessimistic | 0.8 | 2.6 | 6.8 | 84.5 | 70.7 | 52.9 | 34.5 |
| Invasive mechanical ventilation | | | | | | | |
| Evidence-based | 5.0 | 8.9 | 14.7 | 77.3 | 67.3 | 55.8 | 44.0 |
| Non-informative | 12.6 | 18.3 | 25.2 | 66.7 | 57.6 | 48.1 | 38.8 |
| Skeptical | 10.9 | 17.5 | 26.3 | 63.1 | 51.3 | 39.6 | 28.6 |
| Optimistic | 3.1 | 5.8 | 10.2 | 83.3 | 74.6 | 64.1 | 52.6 |
| Pessimistic | 27.5 | 38.3 | 50.2 | 38.0 | 27.2 | 18.2 | 11.3 |

Posterior probabilities of harm and benefit from sensitivity analyses using different priors on the mortality outcome. This table complements Figure 3.

#### Sensitivity analyses using different baseline risks

##### Supplementary Figure 2

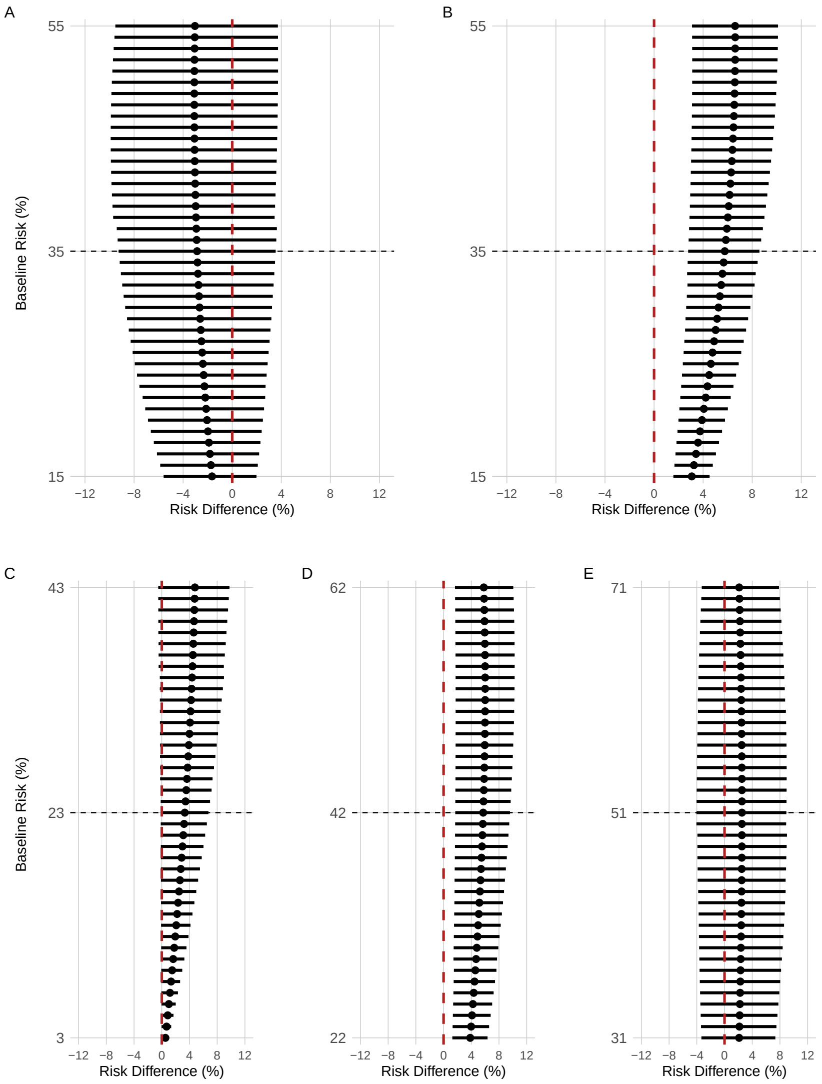

Posterior distributions from sensitivity analyses using different baseline risks on the mortality outcome. Point estimates depict the median and interval bars depict 95% highest density intervals. Each interval bar represents a posterior distribution for the corresponding baseline risk. Horizontal black dashed lines represent the respective baseline risk underlying other analyses (Figure 2), i.e., risk in the control group in the RECOVERY trial for each subgroup (Table 1). Vertical red dashed line represent 0% risk difference. Panel A shows results for patients not using corticosteroids; Panel B shows results for patients using corticosteroids. Panel C shows results for simple oxygen only; Panel D shows results for non-invasive ventilation; Panel E shows results for invasive mechanical ventilation.

#### Results: Hospital discharge outcome

##### RECOVERY and Priors: Number of events and sample size

###### Supplementary Table 5

| | Control | | | Tocilizumab | | |
|---|---|---|---|---|---|---|
| Study | Events | Total | Risk (%) | Events | Total | Risk (%) |
| Not using corticosteroids | | | | | | |
| RECOVERY | 168 | 367 | 46 | 162 | 357 | 45 |
| COVACTA | 35 | 65 | 54 | 124 | 188 | 66 |
| Using corticosteroids | | | | | | |
| RECOVERY | 873 | 1721 | 51 | 987 | 1664 | 59 |
| COVACTA | 36 | 79 | 46 | 42 | 106 | 40 |
| Simple oxygen only | | | | | | |
| RECOVERY | 635 | 933 | 68 | 697 | 935 | 75 |
| COVACTA | 35 | 44 | 80 | 66 | 78 | 85 |
| CORIMUNO-19 | 49 | 67 | 73 | 52 | 63 | 83 |
| Non-invasive ventilation | | | | | | |
| RECOVERY | 362 | 867 | 42 | 401 | 819 | 49 |
| COVACTA | 19 | 39 | 49 | 56 | 94 | 60 |
| Salvarani | 58 | 63 | 92 | 54 | 60 | 90 |
| Invasive mechanical ventilation | | | | | | |
| RECOVERY | 47 | 294 | 16 | 52 | 268 | 19 |
| COVACTA | 13 | 55 | 24 | 35 | 113 | 31 |

Number of events and sample sizes on the hospital discharge outcome. Studies other than RECOVERY represent the data incorporated in the prior distribution for each subgroup.

#### Prior, RECOVERY, and Posterior distributions

##### Supplementary Figure 3

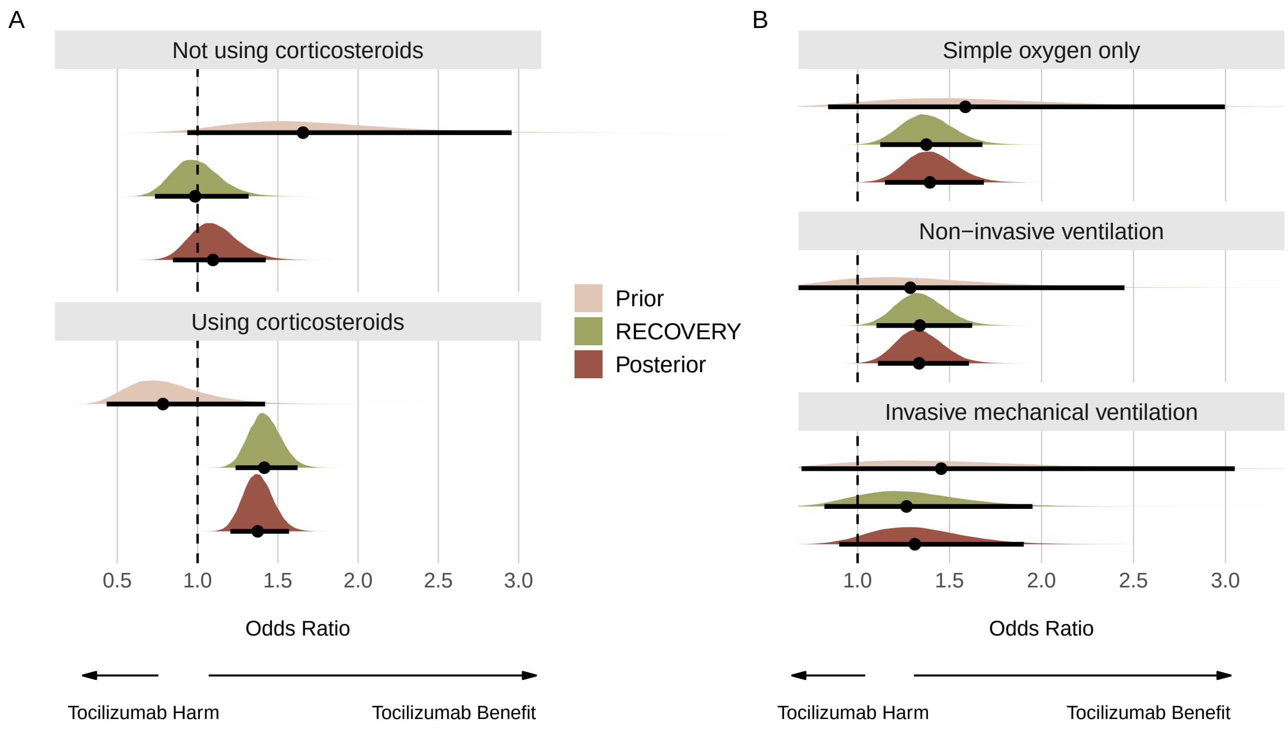

Prior, RECOVERY, and posterior distributions for each subgroup on the hospital discharge outcome. Panel A shows results for subgroups regarding use of corticosteroids. Panel B shows results for subgroups regarding respiratory support. Point estimates depict the median and interval bars depict 95% highest density intervals. Using conjugate normal analyses, these distributions were originally combined in the log-odds ratio scale (Methods section). They were transformed into the odds ratio scale for this figure to aid visual interpretation.

**Supplementary Table 6**

| | 95% Highest Density Interval | | |
|---|---|---|---|
| Distribution | Median | Lower limit | Upper limit |
| Not using corticosteroids | | | |
| Prior | 1.66 | 0.95 | 2.97 |
| RECOVERY | 0.98 | 0.73 | 1.31 |
| Posterior | 1.10 | 0.85 | 1.43 |
| Using corticosteroids | | | |
| Prior | 0.78 | 0.43 | 1.40 |
| RECOVERY | 1.42 | 1.24 | 1.63 |
| Posterior | 1.37 | 1.20 | 1.57 |
| Simple oxygen only | | | |
| Prior | 1.59 | 0.84 | 2.98 |
| RECOVERY | 1.38 | 1.12 | 1.68 |
| Posterior | 1.39 | 1.15 | 1.69 |
| Non-invasive ventilation | | | |
| Prior | 1.29 | 0.68 | 2.47 |
| RECOVERY | 1.34 | 1.10 | 1.62 |
| Posterior | 1.33 | 1.11 | 1.61 |
| Invasive mechanical ventilation | | | |
| Prior | 1.45 | 0.69 | 3.01 |
| RECOVERY | 1.26 | 0.81 | 1.94 |
| Posterior | 1.31 | 0.90 | 1.90 |

95% highest density intervals of evidence-based prior, RECOVERY, and posterior distributions on the hospital discharge outcome. This table complements Supplementary Figure 3.

#### Posterior distribution and probabilities using evidence-based priors

##### Supplementary Figure 4

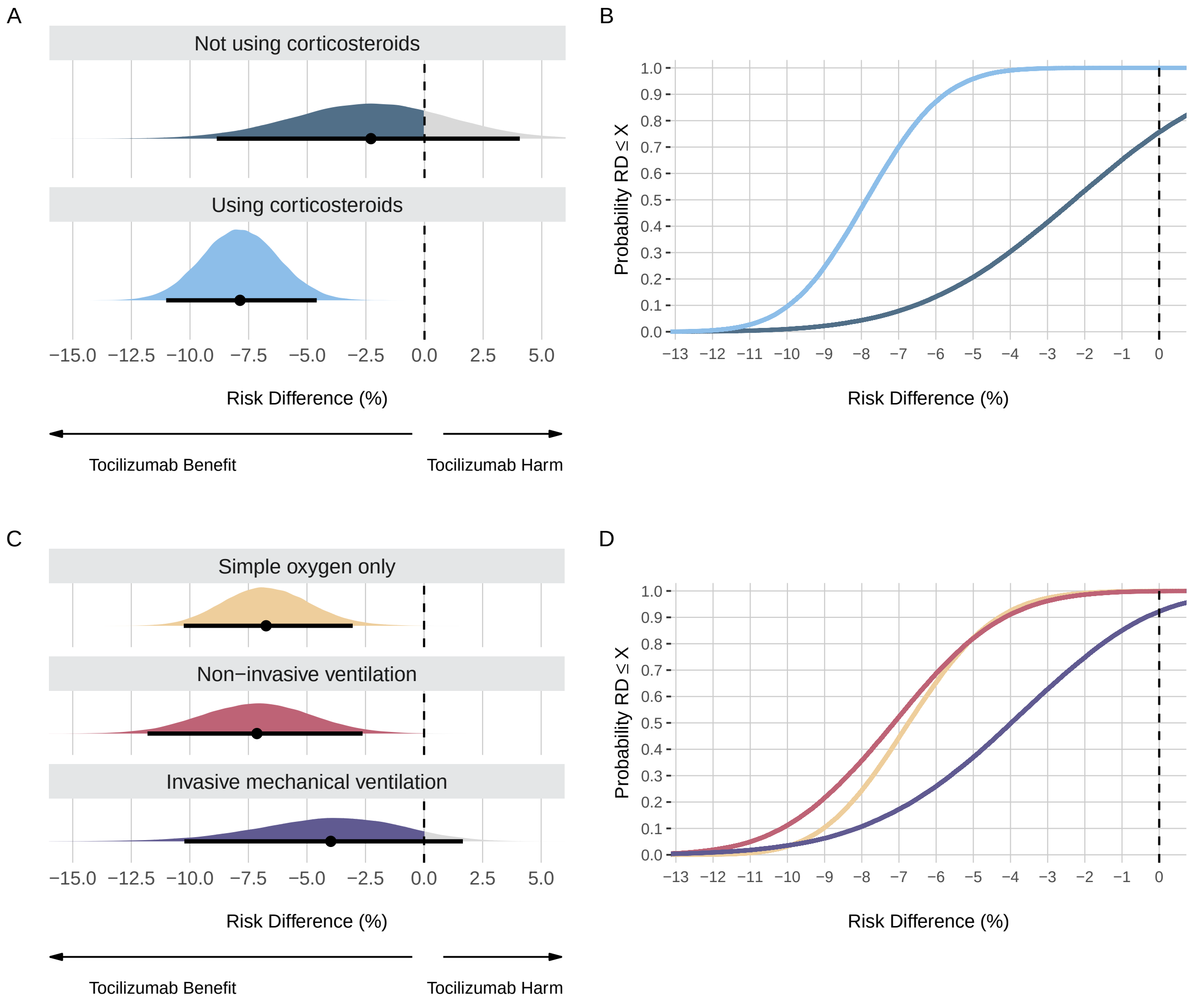

Posterior distributions and probabilities using evidence-based priors on the hospital discharge outcome. Panel A shows the posterior distributions and Panel B shows the cumulative posterior probabilities on subgroups regarding use of corticosteroids. Panel C shows the posterior distributions and Panel D shows the cumulative posterior probabilities on subgroups regarding respiratory support. Panels A and C: Point estimates depict the median and interval bars depict the 95% highest density intervals. Panels B and D: Cumulative posterior distributions correspond to the probabilities that the risk difference (RD) is lower than or equal to the effect size on the X-axis. The colors in Panels B and D match the ones used in Panels A and C.

#### Sensitivity analyses using different priors

##### Supplementary Figure 5

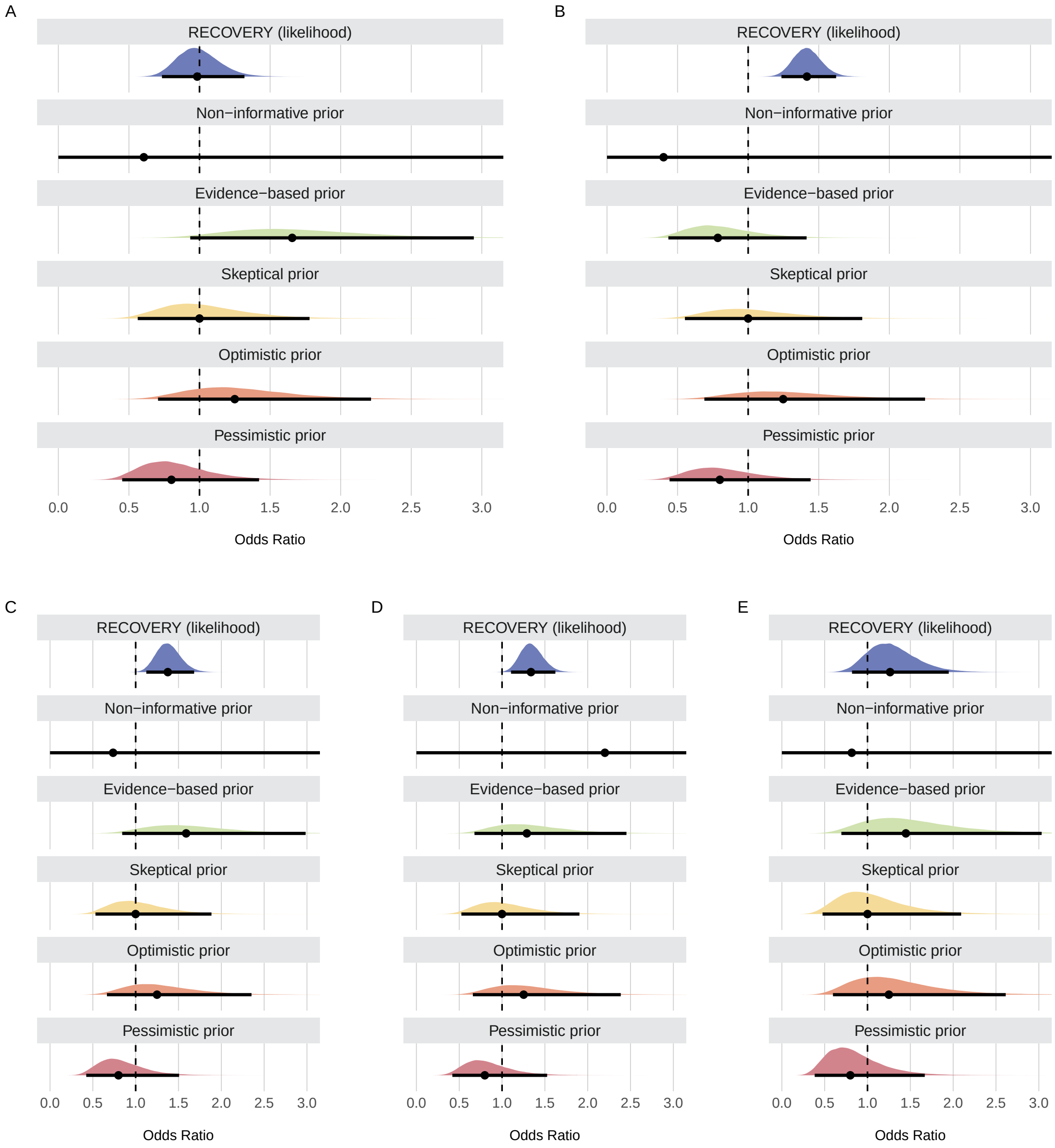

Prior distributions for each subgroup on the hospital discharge outcome. Point estimates depict the median and interval bars depict 95th quantile intervals. Panel A shows results for patients not using corticosteroids; Panel B shows results for patients using corticosteroids. Panel C shows results for simple oxygen only; Panel D shows results for non-invasive ventilation; Panel E shows results for invasive mechanical ventilation.

#### Supplementary Figure 6

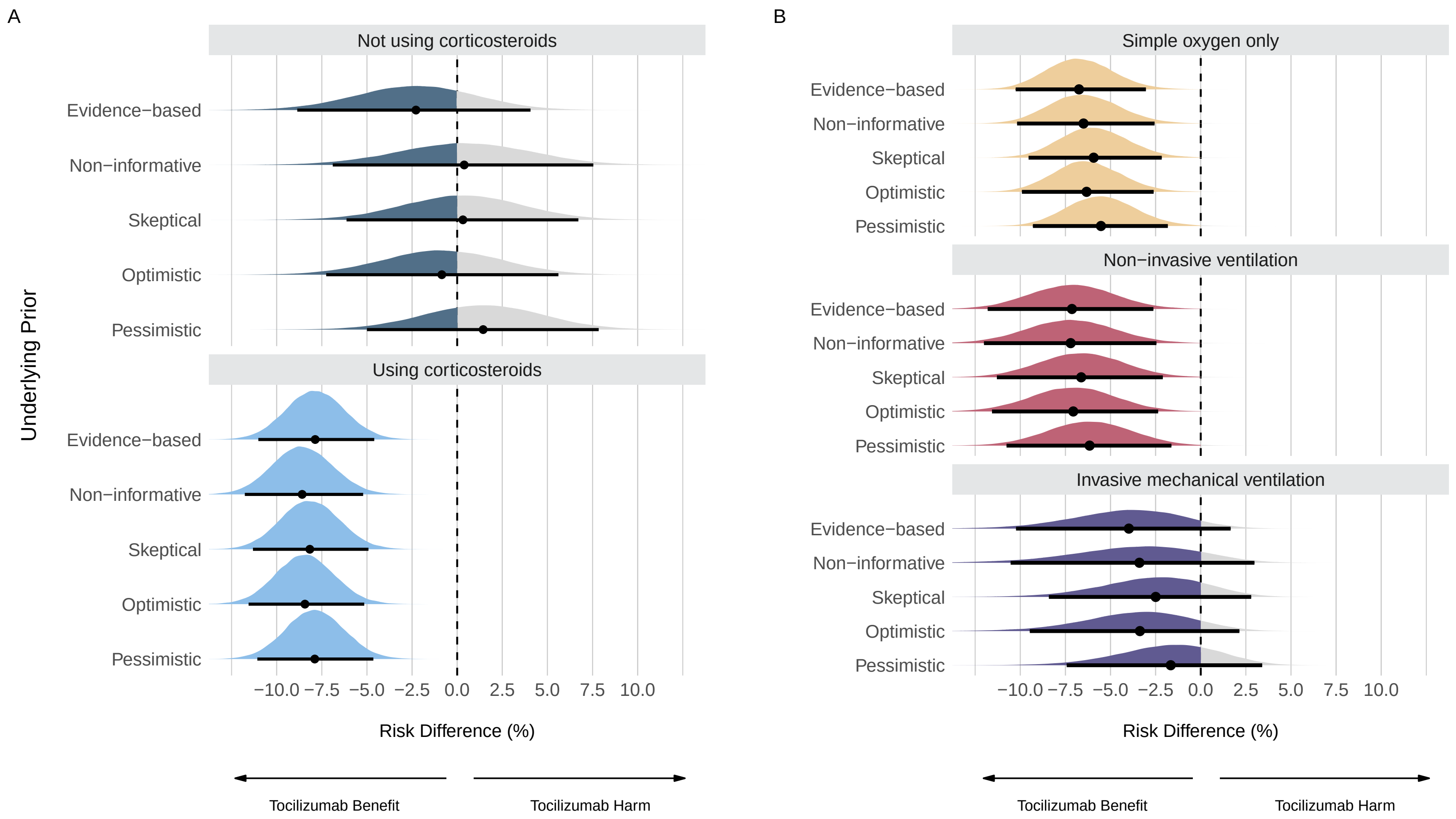

Posterior distributions from sensitivity analyses using different priors on the hospital discharge outcome. Panel A shows posterior distributions on subgroups regarding use of corticosteroids. Panel B shows posterior distributions on subgroups regarding respiratory support. Point estimates depict the median and interval bars depict the 95% highest density intervals.

**Supplementary Table 7**

| Underlying Prior | 95% Highest Density Interval | | |
|---|---|---|---|
| | Median | Lower limit | Upper limit |
| Not using corticosteroids | | | |
| Evidence-based | -2.3 | -8.9 | 4.1 |
| Non-informative | 0.4 | -6.9 | 7.5 |
| Skeptical | 0.3 | -6.1 | 6.7 |
| Optimistic | -0.9 | -7.3 | 5.6 |
| Pessimistic | 1.4 | -5.0 | 7.8 |
| Using corticosteroids | | | |
| Evidence-based | -7.9 | -11.0 | -4.6 |
| Non-informative | -8.6 | -11.8 | -5.2 |
| Skeptical | -8.2 | -11.3 | -4.9 |
| Optimistic | -8.4 | -11.6 | -5.1 |
| Pessimistic | -7.9 | -11.1 | -4.6 |
| Simple oxygen only | | | |
| Evidence-based | -6.7 | -10.3 | -3.0 |
| Non-informative | -6.5 | -10.2 | -2.6 |
| Skeptical | -5.9 | -9.5 | -2.2 |
| Optimistic | -6.3 | -9.9 | -2.6 |
| Pessimistic | -5.5 | -9.3 | -1.8 |
| Non-invasive ventilation | | | |
| Evidence-based | -7.1 | -11.8 | -2.6 |
| Non-informative | -7.2 | -12.0 | -2.5 |
| Skeptical | -6.6 | -11.3 | -2.1 |
| Optimistic | -7.1 | -11.6 | -2.4 |
| Pessimistic | -6.2 | -10.8 | -1.6 |
| Invasive mechanical ventilation | | | |
| Evidence-based | -4.0 | -10.2 | 1.7 |
| Non-informative | -3.4 | -10.5 | 3.0 |
| Skeptical | -2.5 | -8.4 | 2.8 |
| Optimistic | -3.4 | -9.5 | 2.1 |
| Pessimistic | -1.7 | -7.4 | 3.4 |

Median and 95% highest density intervals of posteriors distributions from sensitivity analyses with different priors on the hospital discharge outcome. This table complements Supplementary Figure 6.

**Supplementary Table 8**

| Underlying Prior | Propability of Benefit | | | | Probability of Harm | | |
|---|---|---|---|---|---|---|---|
| | Pr(< -3%) | Pr(< -2%) | Pr(< -1%) | Pr(< 0%) | Pr(> 1%) | Pr(> 2%) | Pr(> 3%) |
| Not using corticosteroids | | | | | | | |
| Evidence-based | 41.5 | 53.4 | 65.1 | 75.6 | 15.9 | 9.6 | 5.3 |
| Non-informative | 17.9 | 25.9 | 35.3 | 45.7 | 43.5 | 33.2 | 24.0 |
| Skeptical | 15.7 | 24.2 | 34.4 | 46.2 | 41.8 | 30.4 | 20.6 |
| Optimistic | 25.5 | 36.3 | 48.2 | 60.1 | 29.0 | 19.5 | 12.1 |
| Pessimistic | 8.9 | 14.8 | 22.9 | 33.1 | 55.4 | 43.2 | 31.7 |
| Using corticosteroids | | | | | | | |
| Evidence-based | 99.8 | 100.0 | 100.0 | 100.0 | 0.0 | 0.0 | 0.0 |
| Non-informative | 100.0 | 100.0 | 100.0 | 100.0 | 0.0 | 0.0 | 0.0 |
| Skeptical | 99.9 | 100.0 | 100.0 | 100.0 | 0.0 | 0.0 | 0.0 |
| Optimistic | 99.9 | 100.0 | 100.0 | 100.0 | 0.0 | 0.0 | 0.0 |
| Pessimistic | 99.8 | 100.0 | 100.0 | 100.0 | 0.0 | 0.0 | 0.0 |
| Simple oxygen only | | | | | | | |
| Evidence-based | 97.3 | 99.2 | 99.8 | 100.0 | 0.0 | 0.0 | 0.0 |
| Non-informative | 95.6 | 98.5 | 99.6 | 99.9 | 0.0 | 0.0 | 0.0 |
| Skeptical | 93.3 | 97.7 | 99.3 | 99.8 | 0.0 | 0.0 | 0.0 |
| Optimistic | 95.6 | 98.6 | 99.6 | 99.9 | 0.0 | 0.0 | 0.0 |
| Pessimistic | 90.2 | 96.2 | 98.8 | 99.7 | 0.1 | 0.0 | 0.0 |
| Non-invasive ventilation | | | | | | | |
| Evidence-based | 96.2 | 98.6 | 99.6 | 99.9 | 0.0 | 0.0 | 0.0 |
| Non-informative | 95.7 | 98.4 | 99.5 | 99.8 | 0.0 | 0.0 | 0.0 |
| Skeptical | 93.9 | 97.6 | 99.2 | 99.8 | 0.1 | 0.0 | 0.0 |
| Optimistic | 95.9 | 98.5 | 99.6 | 99.9 | 0.0 | 0.0 | 0.0 |
| Pessimistic | 91.1 | 96.3 | 98.7 | 99.6 | 0.1 | 0.0 | 0.0 |
| Invasive mechanical ventilation | | | | | | | |
| Evidence-based | 62.8 | 74.8 | 85.0 | 92.2 | 3.5 | 1.2 | 0.4 |
| Non-informative | 54.6 | 66.1 | 76.5 | 85.3 | 8.0 | 3.8 | 1.5 |
| Skeptical | 43.2 | 56.8 | 70.3 | 82.0 | 9.4 | 4.1 | 1.4 |
| Optimistic | 55.0 | 68.3 | 79.8 | 88.7 | 5.4 | 2.0 | 0.6 |
| Pessimistic | 32.0 | 45.2 | 59.5 | 73.2 | 15.5 | 7.5 | 3.0 |

Posterior probabilities from sensitivity analyses using different priors on the hospital discharge outcome. This table complements Supplementary Figure 6.

#### Sensitivity analyses using different baseline risks

##### Supplementary Figure 7

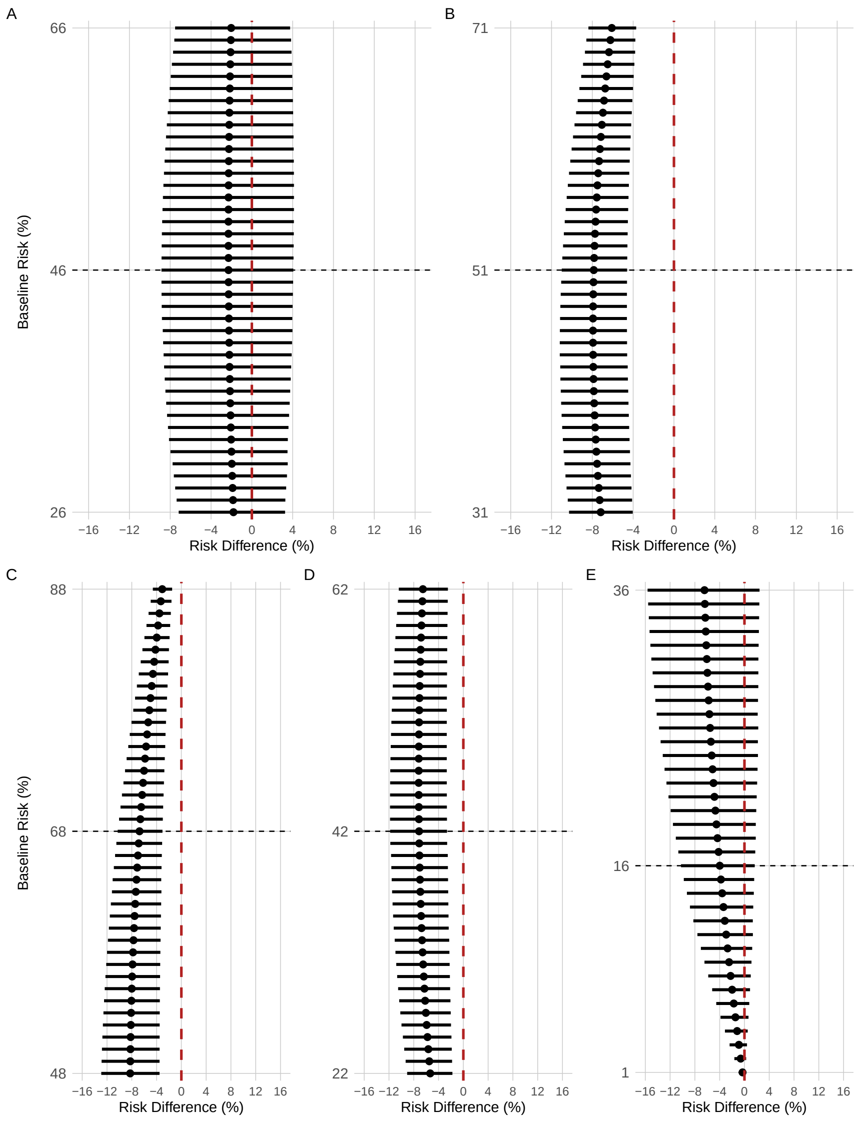

Posterior distributions from sensitivity analyses using multiple different baseline risks. Each interval bar represents a posterior distribution for the corresponding baseline risk. Horizontal black dashed lines represent the respective baseline risk underlying other analyses (Supplementary Figure 4), i.e., risk in the control group in the RECOVERY trial for each subgroup (Supplementary Table 5). Vertical red dashed line represent 0% risk difference. Point estimates depict the median and interval bars depict 95% highest density intervals. Panel A shows results for patients not using corticosteroids; Panel B shows results for patients using corticosteroids. Panel C shows results for simple oxygen only; Panel D shows results for non-invasive ventilation; Panel E shows results for invasive mechanical ventilation.

**Supplementary Figure 8**

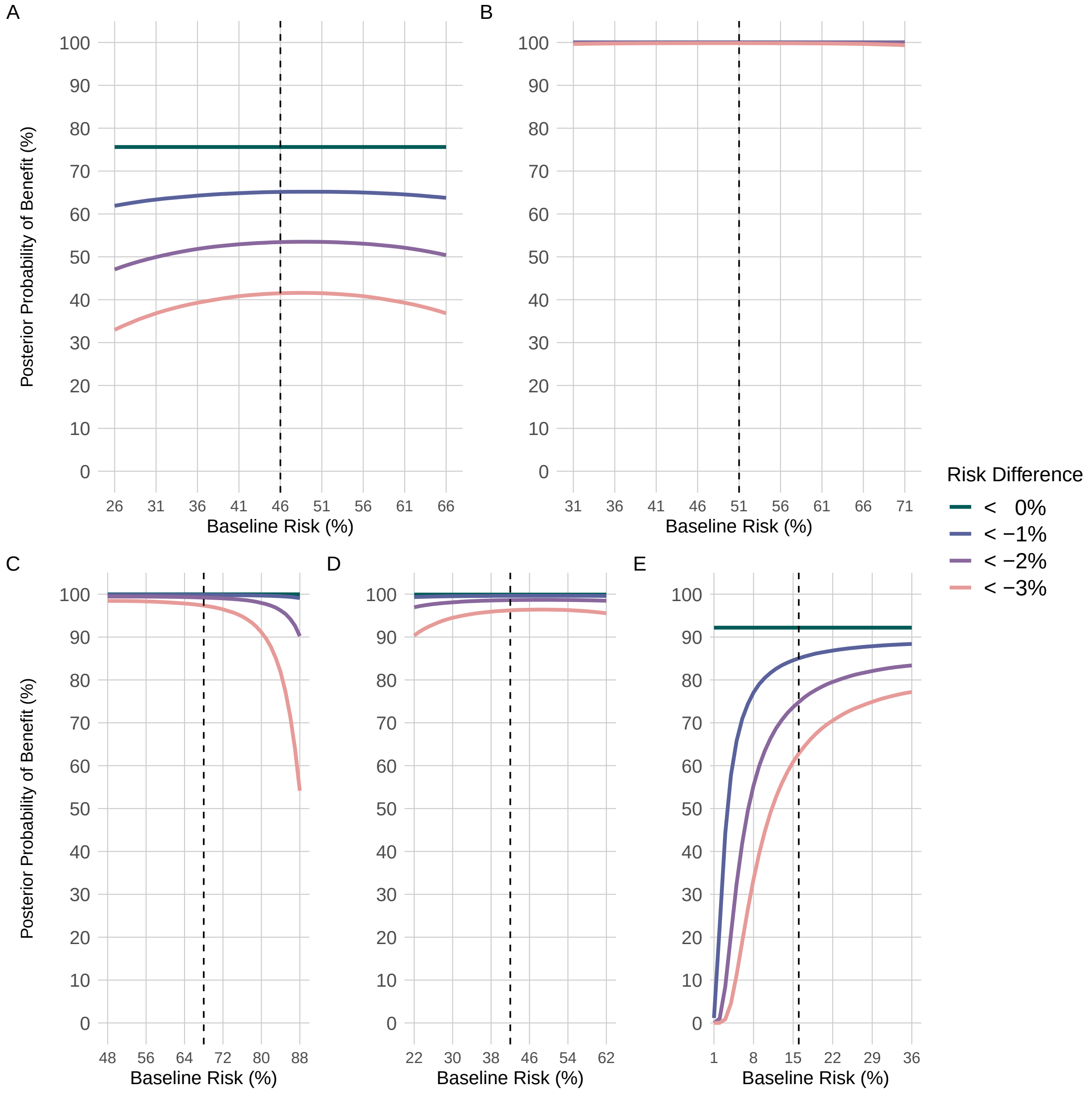

Posterior probabilities from sensitivity analyses using multiple different baseline risks on the hospital discharge outcome. Panel A shows results for patients not using corticosteroids; Panel B shows results for patients using corticosteroids. Panel C shows results for simple oxygen only; Panel D shows results for non-invasive ventilation; Panel E shows results for invasive mechanical ventilation. Each line represents the posterior probability of benefit for a specific cutoff, such as risk difference lower to 0%, 1%, 2% and 3%. Vertical black dashed lines represent the respective baseline risk underlying other analyses (Supplementary Figure 4), i.e., risk in the control group in the RECOVERY trial for each subgroup (Supplementary Table 5).
